# Circadian immunity, sunrise time and the seasonality of respiratory infections

**DOI:** 10.1101/2021.03.29.21254556

**Authors:** Ziyi Mo, Armin Scheben, Joshua Steinberg, Adam Siepel, Robert Martienssen

## Abstract

The innate and adaptive immune response are regulated by biological clocks, and circulating lymphocytes are lowest at sunrise. Accordingly, severity of disease in mouse models is highly dependent on the time of day of viral infection. Here, we explore whether circadian immunity contributes significantly to seasonality of respiratory viruses, including influenza and SARS-CoV-2. Susceptibility-Infection-Recovery-Susceptibility (SIRS) models of influenza and SIRS-derived models of COVID-19 suggest that local sunrise time is a better predictor of the basic reproductive number (*R*_0_) than climate, even when day length is taken into account. Moreover, these models predict a window of susceptibility when local sunrise time corresponds to the morning commute and contact rate is expected to be high. Counterfactual modeling suggests that retaining daylight savings time in the fall would reduce the length of this window, and substantially reduce seasonal waves of respiratory infections.

## Introduction

Severe acute respiratory syndrome coronavirus-2 (SARS-CoV-2) is responsible for the ongoing COVID-19 pandemic. Coronavirus and other respiratory disease outbreaks are often seasonal^1,2^, and the first wave of COVID-19 in the Northern Hemisphere occurred in the spring of 2020, followed by an even more severe second wave in the fall and winter, and an emerging third wave in the spring of 2021. These waves are usually attributed to lower outdoor temperature and humidity during winter which impact survival of free viral particles^1,3^, even though most infections occur indoors^2^. However, seasonal models based on climate-dependence of other coronaviruses and of influenza fail to predict SARS-CoV-2 infection rates, suggesting instead that the supply of susceptibility is the driving factor during the pandemic phase^1,4^.

The 24-hour light-dark cycle entrains “biological clocks” in animals and plants to keep track of circadian time (L. *circa diem* or *around one day*), and to perform diurnal (daily) activities, such as photosynthesis in plants or sleep in mammals. However, day length varies by latitude and by season, so that circadian rhythms must be frequently reset by dark-light transitions. In this way, expression of “morning genes” can accurately anticipate sunrise the following day (which always occurs at ZT0 in Zeitgeber Time: G. *zeit geber*, or *time giver*). In plants, circadian clocks measure day length by illumination of the leaves, and accurately predict the season for flowering. In humans, circadian rhythms are entrained by retinal exposure and innervation of the suprachiasmatic nucleus (SCN) in the hypothalamus^5^. In both cases, light is detected by the conserved cryptochrome blue-light receptor.

In mammals, some of the most prominent targets of clock-encoded regulators are involved in the innate and adaptive immune response^5^, with a nadir in circulating leukocyte counts at sunrise (ZT0)^5,6^. Target genes include many components of the innate immunity system used in viral defence, including toll-like receptors (TLR), interferon receptors, and interleukin-6, all of which have important roles in the SARS-CoV-2 response^5^. Consistently, studies have shown that shift-workers are more susceptible to respiratory viral infections and cardiovascular complications^7^. Mice, on the other hand, are nocturnal, and in a recent study, intranasal infections of mice with influenza virus resulted in 80% survival when infections were performed at ZT23, but only 30% survival at ZT11, the beginning of their active phase^8^. In contrast, mutants in the key clock gene *Bmal1* had similar survival rates when infected at ZT23 and ZT11^8^. Furthermore, in a recent study of free-running circadian monocytes infected with SARS-CoV-2 in culture, both phagocytosis and interleukin activity were massively increased when infection occurred at ZT6 relative to ZT17, relative to clock gene expression maxima^9^.

It has previously been suggested that influenza and many coronaviruses thrive during the winter months because of seasonal variation in innate immunity^7,10^. However, in a recent survey of haematological data from the UK Biobank, circulating monocyte, lymphocyte and neutrophil counts varied 5-10 times more during a single day than they did in spring and winter^6^, with a 25-44% drop when measured at 9am relative to afternoon and evening^5,6^. Regardless of the precise mechanism, we hypothesized that added vulnerability at sunrise could increase the basic reproductive number (*R*_0_) and symptomatic severity of infectious diseases such as influenza and COVID-19. In particular, we hypothesized that *R*_0_ would be elevated when sunrise coincided with high contact rate, which occurs during the morning commute.

## Results

### Application to influenza data

To determine whether the publicly available data for influenza support this hypothesis, we devised several variants of a Susceptibility-Infection-Recovery-Susceptibility (SIRS) model^1,11,12^ and fitted them to a decade’s worth of weekly epidemiological data (ranging from 2010–11 to 2019–20) for individual US states from the US CDC’s FluView report (www.cdc.gov/flu/weekly; see **Supplementary Materials**). Similar to other SIRS models, our model partitions each state’s population into compartments of “susceptible” (*S*), “infected” (*I*), and “recovered” (*R*) individuals, and permits the sizes of these compartments to change over time according to a system of differential equations (see **Supplementary Materials, Supplementary Fig. S2A**). These equations are parameterized by the mean infection period, *D*, the duration of immunity, *L*, and the time-varying basic reproductive number, *R*_0_(*t*).

We defined *R*_0_(*t*), in turn, to be a function of one or more time- and location-varying covariates that could plausibly drive seasonal cycles of influenza. Following previous studies^1,13,14^, we included (absolute) specific humidity (*h*) as a potential climatological covariate and assumed that *R*_0_(*t*) exponentially decays as *h*(*t*) increases. Notably, specific humidity is closely correlated with temperature, relative humidity, solar radiation, and other climatological variables but has been shown to be a major, and likely the predominant, determinant of influenza seasonality^13^. In addition, we included potential covariates for the day length (*d*) and local sunrise time (*s*) associated with each location and time-point in our data set, taking population-weighted averages of these values across states (see **Supplementary Materials**). We assumed Gaussian relationships between *R*_0_(*t*) and *d*(*t*), and between *R*_0_(*t*) and *s*(*t*), with maxima defined by free parameters *d*_0_ and *s*_0_, respectively (**Fig. 1A**). We chose to fix the mean infection period to *D*=5 days and the duration of immunity to *L*=40 weeks, based on previously estimated values for flu (see **Supplementary Materials**); thus, the time-varying dynamics of the SIRS model were entirely determined by the weekly estimates of *R*_0_(*t*), and the only free parameters to be estimated from the data were those that define the relationship between *R*_0_(*t*) and the covariates (*α*_1_, *α*_2_, *α*_3_, *d*_0_, and *s*_0_; see **Fig. 1A**)

**Figure 1.**
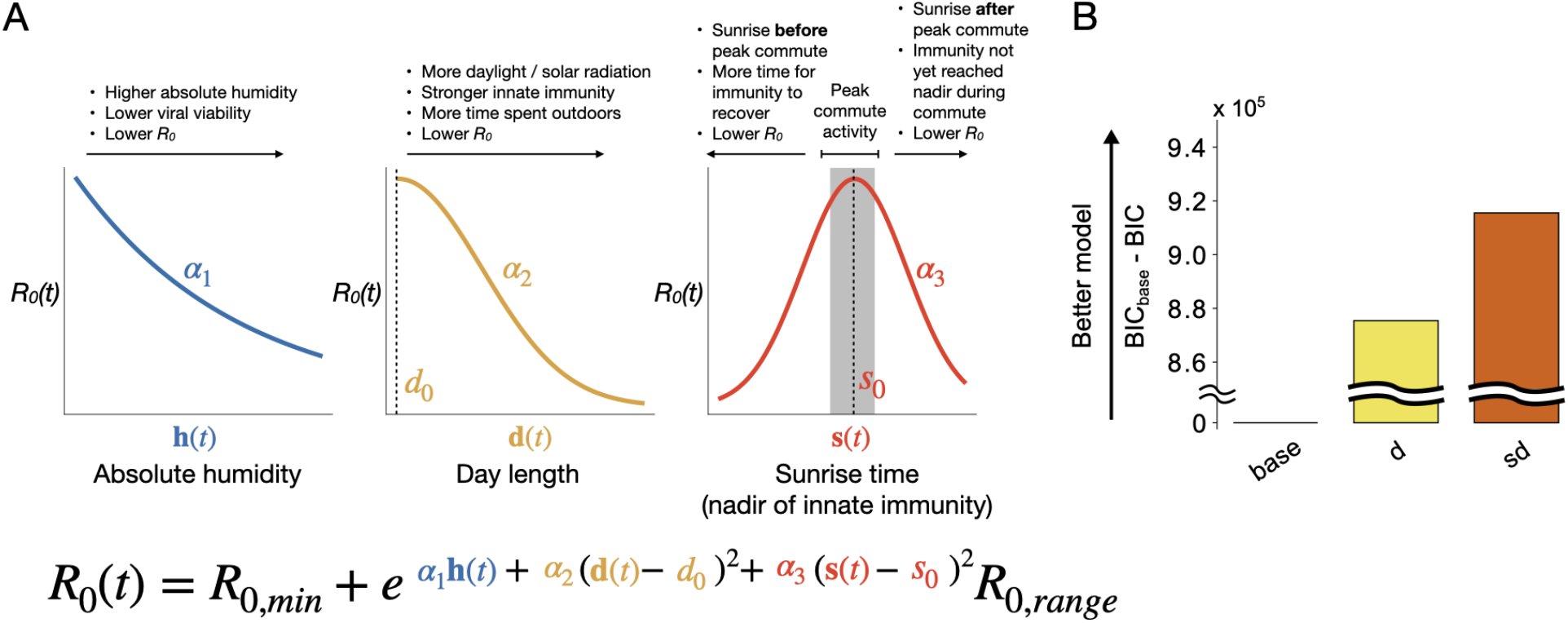
Models for seasonal changes in *R*_0_. **(A)** Functions relating the basic reproductive number *R*_0_ in a Susceptibility-Infection-Recovery-Susceptibility (SIRS) model to (combinations of) time- and location-varying absolute humidity (*h*), day length (*d*) and sunrise time (*s*). Exponential decay is assumed for *h* (with free parameter *α*_1_; see refs. ^1,13^), and Gaussian relationships are assumed for *d* (*α*_2_) and *s* (*α*_3_), with maxima defined by free parameters *d*_0_ and *s*_0_, respectively. *s*_0_ can be interpreted as the peak of the window of susceptibility for infection. The range of *R*_0_ values (parameterized by *R*_0,*min*_ and *R*_0,*range*_) was fixed based on previous estimates for flu models, but fitted to data for COVID-19 models. Notice that *d*_0_ tends to be estimated at values close to zero, making *R*_0_ a monotonically decreasing function of *d* (see **Supplementary Material** for full details). **(B)** SIRS model goodness-of-fit for influenza surveillance data for 40 U.S. states obtained from the US CDC’s FluView report from the 2010–11 season to the 2019–20 season. Goodness-of-fit is measured as the Bayesian Information Criterion (BIC) for the base model minus the BIC for the model in question (and is therefore 0 by definition for “base”). Results are also shown for the day-length (d) and sunrise/day-length (sd) models. The models with humidity (h) showed negligible improvement (**Supplementary Fig. S3**). Model likelihoods were maximized with the penalized likelihood parameter set to *λ*=10, but the relative model fits were similar for other choices of *λ* (see **Supplementary Materials**).

To enable formal comparisons of goodness-of-fit among model variants, we extended the basic SIRS model to not only predict infection rates, but additionally provide a full likelihood for the observed time-series data (**Supplementary Fig. S1**). Briefly, we compute the probability of the reported flu cases under a binomial sampling model, with the sampling probability given by a function of the SIRS-derived infection rate at each time point. To accommodate differences between the general population considered by the SIRS model and the symptomatic subpopulation on which flu tests were performed, we introduced a scaling parameter that is estimated from the data using a penalized likelihood strategy. In this way, we obtained a full generative model for the data and were able to estimate all free parameters by maximum likelihood (ML), with the exception of a single tuning parameter, λ, that controls the strength of the penalty for a truncated sampling probability. We find that the results of our analysis are qualitatively similar for a wide range of choices of λ (**Supplementary Materials**). To verify that the model was implemented appropriately, we compared the version with humidity as the only covariate to a previously published SIRS model for influenza^13^ and found that it performed similarly, modulo some differences in data sets and implementation details (see **Supplementary Materials**).

We evaluated the relative importance of the humidity, day length, and local-sunrise-time covariates by comparing several alternative versions of the model, measuring goodness-of-fit using the Bayesian Information Criterion (BIC) to account for differences in numbers of free parameters. We fitted each version of the model jointly to the 10-year time series of weekly infections for 40 US states, excluding 10 states for which data were limited (AK, FL, ID, ME, NJ, NH, NV, RI, VT, and WY; see **Supplementary Materials**). In each case, the observed covariate values differed among states, but their relationship to *R*_0_(*t*) was defined by free parameters shared across all states. For comparison, we also considered a baseline model that allowed for seasonal variation in *R*_0_(*t*) through a simple sinusoidal function, independent of any environmental covariates.

We found that adding day length as a covariate substantially improved the fit of the baseline model, and adding local sunrise time improved it further by a smaller, but clearly significant, margin (**Fig. 1B**). Interestingly, however, humidity had little benefit in this setting, with the parameter *α*_1_ tending toward zero and versions of the model that included humidity fitting about as well as matched versions without it (**Supplementary Table S1, Fig. S3**). Notably, the improvement in fit from day length could as well represent contributions from any other correlated seasonal variable, such as temperature, solar radiation, or perhaps, time spent outdoors^15^. The additional improvement in fit from local sunrise time, however, must represent a specific impact from this variable, because it is decoupled from day length only by the convention of seasonal daylight savings time. In other words, our model comparison is conservative in attributing as many seasonal effects as possible to day length, and then only attributing to sunrise specific effects that become apparent through the disruption of changing the clocks in the spring and fall. Interestingly, we find that the maximum-likelihood estimate (MLE) of the parameter describing the peak impact of sunrise time on *R*_0_(*t*) occurs between 6 and 7 am (0.5 < *s*_0_ < 0.58; **Supplementary Table S1**), consistent with the typical morning rush hour in most US cities (see **Discussion**). Thus we find that local sunrise time is significantly associated with influenza even after accounting for other seasonal effects, and the nature of this association is consistent with an elevated infection rate when sunrise occurs around the time of the morning commute.

The apparent importance of local sunrise time after accounting for day length suggests a possible counterfactual experiment: by how much could influenza infections be reduced if seasonal daylight savings time were eliminated? Our framework allows this question to be addressed by running our model forward under hypothetical alternative relationships between sunrise time and day length, consistent with various policies for clock time. We focused on three possible policies: (1) the one currently used in most US states (*seasonal DST*); (2) a policy where DST is completely eliminated (*permanent standard time*); and (3) a policy where DST is maintained throughout the year (*permanent DST*). We evaluated all three schemes using average values of the covariates over the past 10 years under the version of the model that allows for both day length and sunrise time to obtain an estimate of the expected effect of each scheme on flu infections, holding all parameters fixed at their MLEs. We found that eliminating seasonal clock changes, through either the permanent standard time or permanent DST policies, did indeed reduce the expected number of annual flu infections (**Fig. 2B**). Interestingly, however, the permanent-DST strategy results in a clearer reduction in infections (6.0%±1.7%) than the permanent-standard-time strategy (0.8%±2.2%) (**Fig. 2B&C**; see **Discussion**). Here, we report model predictions and uncertainty as MLE ± SD, where SD is calculated by bootstrapping over states (see **Discussion** and **Supplementary Materials**). Owing to a variety of factors, this reduction varies considerably across states (**Fig. 2C**), but the permanent DST policy always reduces infections, and always by more than permanent standard time. Thus, our model suggests that eliminating seasonal clock changes could significantly reduce the incidence of seasonal influenza, particularly if permanent DST were adopted (see **Discussion**).

**Figure 2.**
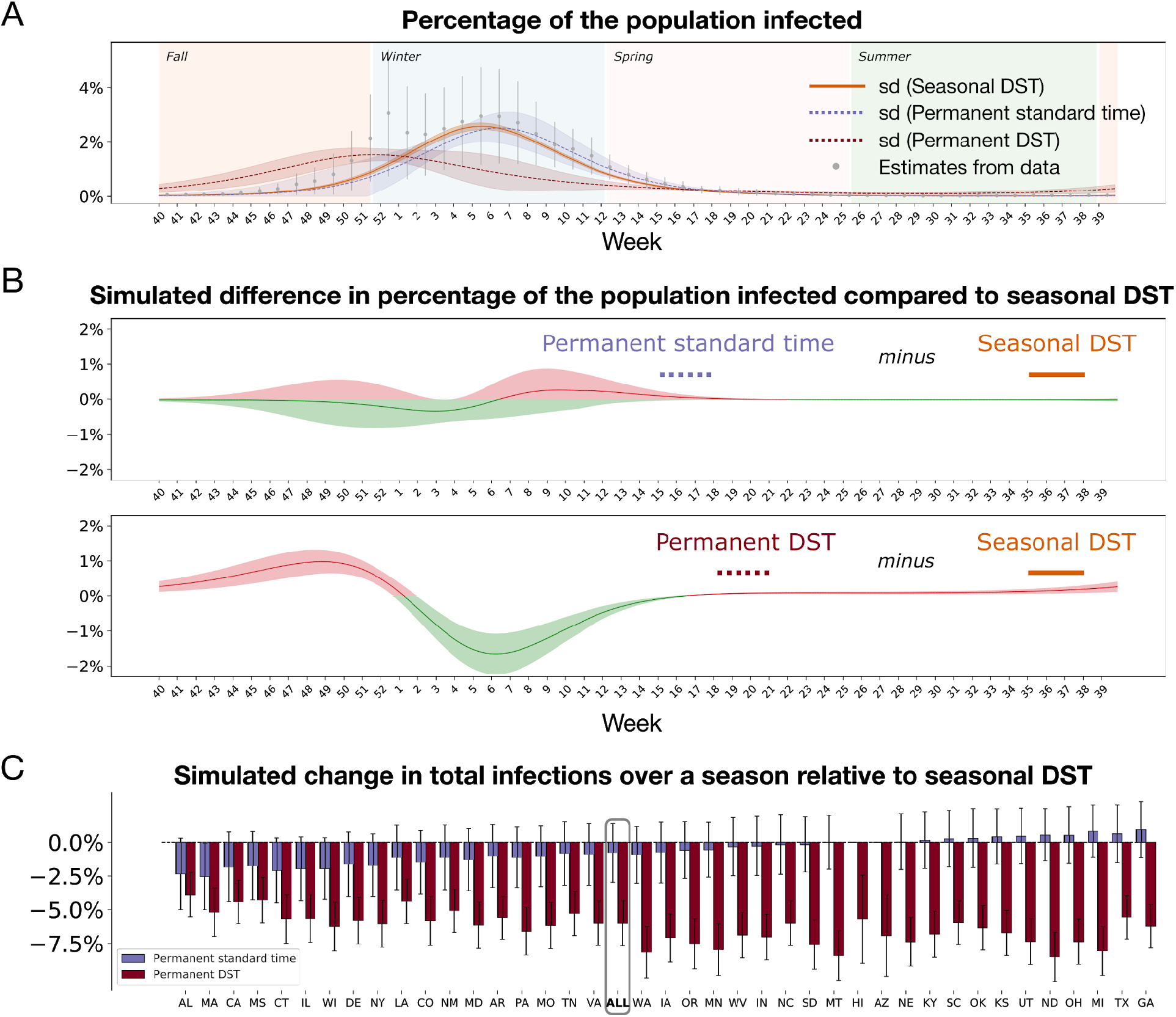
Yearly incidence of seasonal flu under alternative policies for clock time. **(A)** Estimates of the percentage of the population infected across 40 U.S. states over a typical flu season under the sunrise/day-length (sd) model. Results are shown for the fitted model (“Seasonal DST”; based on *λ*=10) as well as the “Permanent standard time” and “Permanent DST” alternatives. For comparison, a summary of the raw data over 10 flu seasons is shown, with error bars indicating one standard deviation. Data are from the US CDC’s FluView report from 2010–11 to 2019–20. (**B**) Reduction (*green*) or increase (*red*) in infections under the Permanent standard time and Permanent DST policies. These plots represent the differences between the corresponding curves in panel **A**. (**C**) Difference in the total number of seasonal flu infections (defined as in **B**) in individual US states. Error bands around the trajectories in **(A), (B)** and error bars in **(C)** indicate one standard deviation based on 50 bootstrap replicates, where the states are resampled in each replicate (see **Supplementary Materials** for details).

### Application to COVID-19 data

We sought next to carry out a parallel analysis of data for COVID-19—to ask whether local sunrise time also helps to explain COVID-19 infection rates, and if so, whether elimination of seasonal clock changes could reduce them. To this end, we aimed to model *R*_0_(*t*) of COVID-19 the same way as with flu, as a function of the population-weighted averages of the same set of time-varying covariates. However, we faced additional technical challenges in this case. First, the data on COVID-19 infections are less reliable than for influenza infections, because of the prevalence of asymptomatic cases, the limited testing capacity, and overlap in symptoms with flu and other respiratory diseases. As a result, we turned to data on deaths and hospitalizations from the COVID Tracking Project (covidtracking.com/data/download/all-states-history.csv), which we expect to be more reliable. To accommodate deaths, we extended the model to include not only the *S-I-R* compartments, but also a “resolving” (*G*) compartment to represent the pool of patients that have exited the stage of initial infection (*I*) and are awaiting resolution as a death (at a rate *δ*) or survival (**Supplementary Fig. S2B**). We also introduced an “exposed” (*E*) compartment between *S* and *I* to allow for the lag due to incubation of SARS-CoV-2 (see ref.^16^ for comparison). To accommodate hospitalization data, we similarly introduced a compartment for hospitalized (*H*) patients (in place of *G*), which infected individuals enter at a hospitalization rate *η* and remain in for a mean time of *κ* (**Supplementary Fig. S2C**, also see ref.^17^). We adapted our sampling models to account for these data, again allowing for a full likelihood approach. Drawing from a variety of sources (see **Supplementary Materials**), we fixed the the death rate at *δ*=1%, hospitalization rate at *η*=2.6%, incubation period at *σ*=5 days, infection period at *γ*=5 days, and mean hospitalization time at *κ*=10 days.

A second major complication with COVID-19 is that, as a newly emerging pandemic, its early stages are particularly difficult to model. We faced two major issues here: first, the “clock” for the pandemic effectively started at different times in different locations, as the virus spread across the US and the world; and second, the data on hospitalizations and deaths from early 2020 are particularly inconsistent and unreliable. To address the first problem, we treated the initial fraction of the population attributed to the “infected” compartment, *I/N*, as a free parameter to be separately estimated for each state. To address the second, we considered only data from on or after March 13, 2020, the date on which the US COVID-19 National Emergency was declared. In an attempt to provide further flexibility across states, we allowed the time of peak local sunrise effect, *s*_0_, to vary across regions (see **Supplementary Materials**). All other parameters were shared across states.

As with influenza, we fitted various versions of these models to U.S. state-level COVID-19 data from March 13, 2020 (or when they were first available, if later) to January 31, 2021, and compared them using the BIC. The results for both the hospitalization and death versions of the model were qualitatively similar to one another, and to the results for influenza. Again, we found that the inclusion of the day-length covariate improved substantially on the baseline model, and the additional inclusion of local sunrise time improved model fit by a smaller, but significant, margin (**Fig. 3A**; **Supplementary Fig. S4**). Again, humidity provided no improvement to versions of the model that already included day length and sunrise (**Supplementary Tables S2 & S5**; **Supplementary Fig. S4**). The version of the model with both day length and local sunrise time showed a reasonably good fit to the state-wise and regional data (**Figs. 3C-F**; **Supplementary Figs. S7-S9**), given the complexities of modeling a newly emerging pandemic. Moreover, our relative model fits and parameter estimates were reasonably robust to different hospitalization and death rates (**Supplementary Fig. S4, Supplementary Tables S3-S7**). The proportion of the population that had been infected and recovered, *R*/*N*, showed some sensitivity to these rates, but, overall, exhibited reasonable agreement with recent surveys of serological prevalence of COVID-19 antibodies^18,19^ (**Supplementary Fig. S5**). The main deficiency of our models was that, while they were able to capture the spring and fall peaks in 2020 COVID cases, they were not able to resolve the spring and summer waves in certain regions of the US, particularly in the south, perhaps because the distinct summer peak reflects violations of our modeling assumptions (see **Discussion**).

**Figure 3.**
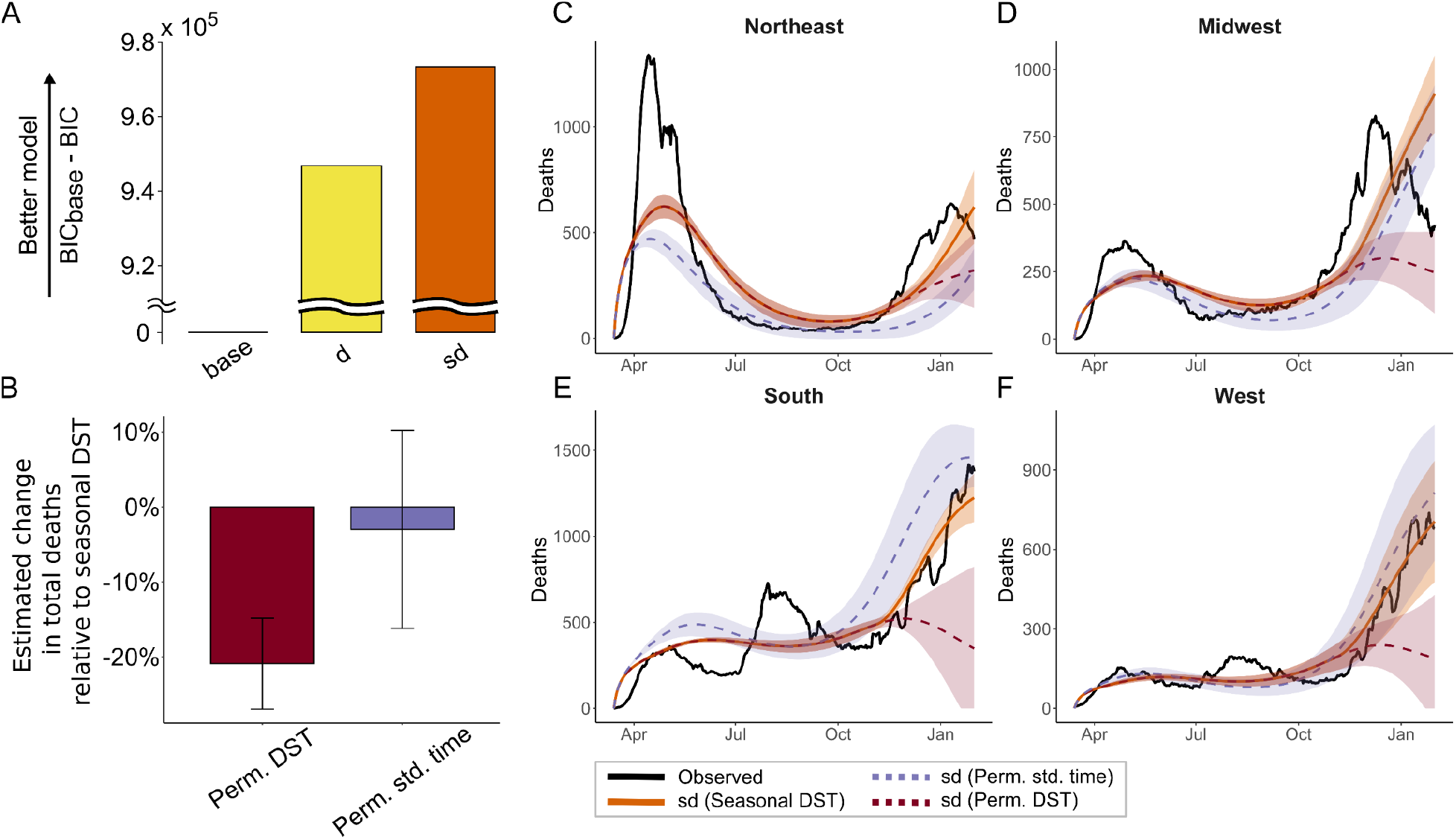
SIRS-derived models fitted to state-level COVID-19 death data for U.S. states. (**A**) Goodness-of-fit of the base, day-length (d), and sunrise/day-length (sd) models, as in **Fig. 1B. (B)** Estimated changes in the total number of COVID-19 deaths between 03/13/2020 and 01/31/2021 under alternative time policies. (**C–F**) Observed COVID-19 death data from the COVID Tracking Project from 03/13/2020 to 01/31/2021 (black) is shown alongside the sunrise/day-length (sd) model fit (solid orange) and projections under alternative time policies (dashed lines). Model fitting and simulations were performed at the state level, and results were subsequently aggregated by census region. Error bands around trajectories in **(C–F)** and error bars in **(B)** indicate one standard deviation based on 91 bootstrap replicates. Model likelihoods were maximized under a death rate of *δ*=1% (see **Supplementary Materials** for full details).

Next, we asked, as with flu, how much COVID-19 incidence might be reduced by eliminating seasonal clock changes. Again, we ran our model forward under three hypothetical time policies, using the day-length and local sunrise time covariates and keeping all parameters fixed at their MLEs. We found, similar to flu, that abandoning seasonal clock changes resulted in considerable reductions in both deaths and hospitalizations (**Fig. 3B, Supplementary Fig. S6**). Remarkably, our models based on *δ*=1% and *η*=2.6% indicated that COVID deaths and hospitalizations over the period from Mar 13, 2020 to Jan 31, 2021 could have been reduced by as much as 20.9% ± 6.1% and 20.7% ± 6.2%, respectively, by the permanent DST policy. Importantly, the relative effects of the permanent-DST and permanent-standard-time policies exhibited considerable state-to-state variation (**Supplementary Fig. S6**). In general, permanent DST offers a larger reduction of the fall and winter peak in the South and West, whereas permanent standard time provides the most advantage in the Northeast, by reducing and shortening the spring peak (**Figs. 3C-F**).

Finally, we extended our analyses to 31 cities from around the world, fitting the sunrise-and-day-length (sd) version of our model to city-level data. The data sets for individual cities are smaller and potentially more reflective of local anomalies, but, importantly, the day length and sunrise time are much more precise at the city level, and therefore may provide more explanatory power. We found fair correlations of our model fit to the observed data across cities (Pearson’s *r*=0.66), with higher correlations in the cities with peak daily deaths exceeding 150, which tended to drive the joint model fitting (*r*=0.78). On the other hand, our bootstrapping analysis revealed higher levels of uncertainty in total predicted deaths for all cities (±18.2%) than for all US states (±4.9%), with particularly high variance in cities with low death counts likely driven by local anomalies such as Melbourne (**Supplementary Fig. S10**). Overall, our model captured epidemiological trends in a range of cities in the Northern and Southern Hemispheres, reiterating our other predictions that seasonal time changes substantially impact infection rates (**Fig. 4**).

**Figure 4.**
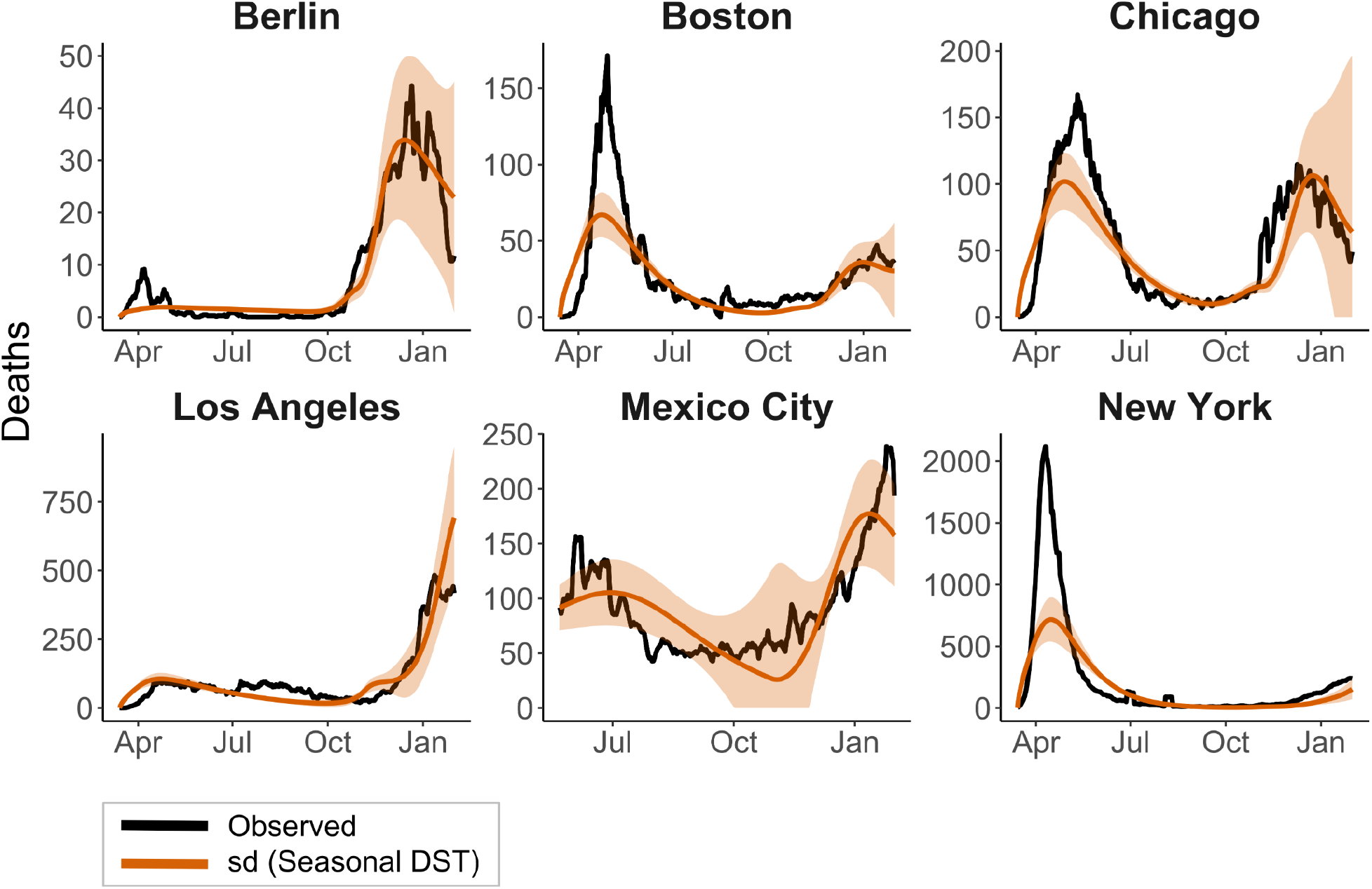
Application of the sunrise/day-length (sd) model to six urban centers around the world. Error bands around trajectories indicate one standard deviation of simulation outcomes based on 47 bootstrap replicates. Raw data are shown in black. Model likelihoods were maximized under a death rate of *δ*=1% (see **Supplementary Materials** for full details).

## Discussion

Following the general Susceptibility-Infection-Recovery-Susceptibility (SIRS) paradigm^1,11,16,17^, we developed models for both influenza and COVID-19, and fitted them to available public data for infections (in influenza) and hospitalizations and deaths (in COVID-19). We adapted these models to capture the relationship between the basic reproductive number *R*_0_ and several key covariates—including humidity, day length, and local sunrise time—allowing *R*_0_ to change in a seasonal manner as a function of the covariates. Despite some variability across states and cities, we found, overall, that these models fit the data remarkably well, especially given the challenges stemming from heterogeneity in data sources and reporting standards, the emergence of new strains of both viruses, and the many challenges of adequately modeling the early stages of the COVID-19 pandemic.

We found, in particular, that day length is a good predictor of *R*_0_ for both influenza and COVID-19, suggesting that it plays a major role in seasonal variation in these diseases. Interestingly, humidity had little effect in the context of the other covariates, resulting in negligible improvements once day length and/or local sunrise time were accounted for, despite that it appears to be one of the best available climatological covariates^1,13^. As noted, our approach does not allow us to distinguish precisely between day length and a variety of other covariates that are highly correlated with it, such as temperature, solar radiation, time spent outdoors, or other variables that could make significant contributions to seasonal variation^2,7^. Our test of local sunrise time, however, is conservative, because this variable is almost perfectly correlated with day length except for the perturbation of seasonal daylight savings time. Thus, the significantly improved fit to the data that we observe when we add the sunrise time covariate to a day-length model can be obtained only because the model exploits this perturbation.

Owing to the close correlation with day length, it is likely that our modeling approach underestimates the importance of local sunrise time; that is, our conservative strategy likely misattributes a portion of the sunrise effect to day length. Indeed, a number of more circumstantial observations suggest an important contribution from local sunrise time. For example, the first wave of SARS-CoV-2 infections occurred around the equinox, so that local sunrise time occurred within a one-hour window at the incidence peak of daily cases in cities across the globe (**Supplementary Fig. S11**). This sunrise-peak co-incidence occurred despite wide variation in case load, reporting rates, fatality rates, latitude and calendar date. Interestingly, most equatorial countries, where sunrise always occurs in the same window, have either largely escaped the pandemic or else have ongoing outbreaks without an obvious peak, presumably reflecting a near-constant *R*_0_ throughout the year. As our model shows, subsequent waves are generally limited by reductions in the susceptible population (*S*). Strikingly, however, the third wave currently spreading in Europe and the US in 2021 has similar patterns of emergence as those observed in early 2020, likely reflecting a predictable seasonal trend.

Why would sunrise time so significantly impact the reproductive number? A likely explanation is that reduced circadian immunity at sunrise contributes substantially to an elevated *R*_0_ when it occurs during the morning commute, when the contact rate is high. Indeed, the estimated *s*_0_ parameter, indicating the peak impact of sunrise time on *R*_0_, suggests that the window of susceptibility that we have identified corresponds reasonably well to commuting hours at most locations examined. These modeling results are concordant with the recent observation of considerably greater diurnal than seasonal variation in leukocyte counts in the UK Biobank^6^.

Of course, social distancing measures, shutdowns, national holidays and travel restrictions also greatly impact *R*_0_, and have complex relationships to day length, for example. However, the sunrise model leads to some clear predictions. First, workers who start at dawn, such as agricultural and outdoor market workers, would be particularly vulnerable, supporting a recent proposal to change working hours to take circadian immunity into account^20^. Second, and more broadly, the model suggests a simple intervention to reduce the length and severity of “winter” epidemics: abandoning the practice of changing the clocks each spring and fall. As the post-pandemic trajectory of COVID-19 hinges on a complex interplay between immunity and seasonality^1,21–23^, a change in clock policy could serve as a relatively inexpensive mitigation strategy in conjunction with vaccination efforts and other interventions.

We attempted to quantify the potential effect of this intervention by running our models forward under the alternative time policies. The predicted reductions in overall influenza cases under permanent DST were substantial, ranging from 4.3–7.7% (including estimated standard errors; see **Supplementary Materials**). Even more strikingly, we predicted an overall reduction in COVID-19 deaths of 14.8–27.0% and an overall reduction in hospitalizations of 14.5–26.9%. Notably, however, the COVID-19 estimates exhibited much greater variance, presumably due to the quality and heterogeneity of the COVID-19 epidemiological data. Interestingly, our models suggest—for both influenza and COVID-19—that the permanent-DST strategy is considerably more effective at reducing infections, overall, than the permanent-standard-time strategy. The reason for this difference is that permanent DST tends to lead to a substantially earlier and milder winter peak, whereas permanent standard time tends to delay the winter peak only slightly. These effects are particularly clear in the case of flu, where we have more data and closer-to-equilibrium conditions (**Fig. 2A&B**). They can be understood by considering the timing and duration of the window of susceptibility during which the typical morning commute coincides with local sunrise time, and hence, increased susceptibility. Seasonal DST tends to artificially extend this window, even doubling it in some locations (**Fig. 5**), thereby contributing to a pronounced increase in infections. Of the two strategies that do not involve changing the clocks, permanent standard time causes this window to occur substantially earlier in the fall, which has the effect of not only making infections peak earlier but also considerably reducing their number. In some states (such as GA; see **Fig. 5**), permanent standard time causes the window of susceptibility to occur in the summer or early fall, when infections are otherwise more containable.

**Figure 5.**
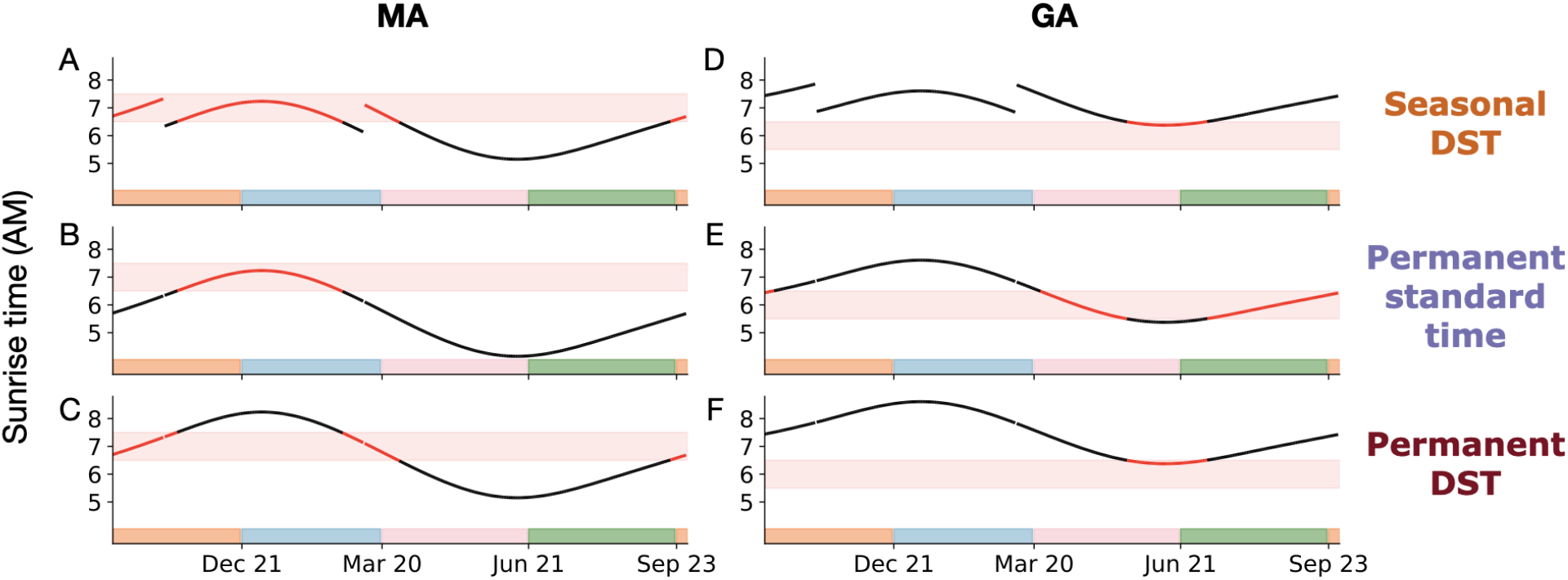
Illustration of impact of time policy on window of susceptibility for infection. The curves represent population-density-weighted local sunrise time in Massachusetts (**A–C**) and Georgia (**D–F**), under the seasonal DST (**A&D**), permanent standard time (**B&E**), and permanent DST (**C&F**) policies. The red horizontal band in each panel indicates the approximate window of susceptibility in each location (defined by the estimated *s*_*0*_ parameter in the sunrise/day-length (sd) model; **Supplementary Tables S1 & S4**). Colors on the *x*-axis indicate the four seasons. Notice that seasonal DST substantially prolongs the interval where local sunrise time coincides with the window of susceptibility in Massachusetts. Changing to permanent daylight savings time would push sunrise one hour later in the late fall and winter **(C&F)**, ensuring that innate immunity does not reach its nadir until after the window of susceptibility. Conversely, changing to permanent standard time would shift sunrise one hour earlier in the spring, summer, and early fall. In Massachusetts, this would increase the interval between sunrise and the window of susceptibility, and allow circadian innate immunity to recover after the low point at dawn **(B)**. In Georgia, however, sunrise occurs too late in the summer months for this improvement to be realized.

A major challenge in this exercise is quantifying the uncertainty in these counterfactual predictions. We attempted to account for uncertainty by bootstrap-resampling at the level of the states (see **Supplementary Materials** and ref.^12^). This bootstrapping approach accommodates various sources of error, including failures in numerical optimization or selection of suboptimal local maxima, and leads to substantial estimates in uncertainty, with the range of counterfactual predictions by our COVID-19 models often exceeding 10 percentage points (**Fig. 3B, Supplementary Fig. S6**). However, this approach still has the limitation that it only accounts for uncertainty in parameter values, and not for misspecification of the model itself—for example, owing to changes in hospitalization or death rates over time, evolving mitigation strategies, or the omission of critical covariates. For these reasons, it is likely that our error bars underestimate the true uncertainty in our predictions, especially for COVID-19.

Some observations did suggest that our modeling assumptions are not adequate in all respects. For example, while our models were able to capture the spring and fall peaks in 2020 COVID-19 cases, they were not able to resolve the spring and summer waves in certain regions of the US, particularly in the south. This distinct summer peak may reflect evolving mitigation strategies during the first half of 2020, seasonal day-break hours for vulnerable groups such as agricultural workers, or other missing covariates in our model. In addition, the versions of the model we fitted to hospitalization and death data, while generally concordant, did not agree in all respects. For example, the estimated parameters differ somewhat between the two versions (**Supplementary Tables S2 & S5**), possibly owing to deviations from our assumption of constant death and hospitalization rates. Similarly, we obtained somewhat different parameter estimates at the city and state levels. One recurring issue is that our estimation procedure inevitably sacrifices goodness of fit for cities or states with smaller numbers in order to accommodate ones with larger numbers, causing population centers to dominate the model fitting procedure. These limitations of our models attest to the substantial impact of lockdown and social distancing measures on the epidemiology of COVID-19, and therefore their indispensable role in combating pandemic respiratory viral diseases^23–27^. Nevertheless, despite these caveats, the models generally fit the data fairly well and were in reasonable agreement with one another and with other published observations (**Supplementary Figs. S5, S7-S10**). Importantly, the effect of sunrise, even by our conservative estimation strategy, appears to be substantial, it remains so under resampling approaches and alternative choices of the hospitalization and death rates, and it is present even when additional environmental covariates (such as humidity) are included. Therefore, we believe the evidence is strong that a change in clock policy would have a real impact in reducing infection rates of both influenza and COVID-19, and likely for other respiratory infections as well.

The H1N1 influenza pandemic of 1918-1920 peaked in multiple spring and fall waves, the most-deadly being the second wave that began in September and October of 1918^28^. DST was first introduced in World War I, coinciding in the United States with the first and second waves (March–November 1918). The third wave in the spring of 1919 was less severe and occurred after DST was widely abandoned. DST was re-adopted as a Federal standard in 1967, just in time for the H3N2 Hong Kong Flu (1968–70), and was expanded in 2007, just in time for H1N1 swine flu in 2009. DST was also observed in Hong Kong during the Asian (1957) and Hong Kong (1968) flu pandemics. But currently, Asian and African countries do not have DST, and have lower incidence and severity of COVID-19 than most countries in the Americas, Europe and the Middle East, where seasonal clock changes are widely observed (**Supplementary Fig. S12**). While there are many important exceptions, and many confounding factors, this worldwide trend supports our counterfactual modelling predictions that seasonal clock changes should be abandoned, a proposal already approved (though not yet implemented) in the European Union^29^. If *zeitgeber* zero hour—sunrise—does play a role in transmission and severity, these proposals to abandon seasonal clock changes are likely to reduce the impact of circadian immunity on ongoing and future seasonal epidemics.

## Data Availability

All data and code used in this study are publicly available (github.com/ziyimo/circ-immu).

## Acknowledgements

This research was supported, in part, by US National Institutes of Health grant R35-GM127070 (to A.S.), the Louis Morin Charitable Trust (J.S.), the Gladys & Roland Harriman Fellowship (Z.M.), the Howard Hughes Medical Institute (R.M.), and the Simons Center for Quantitative Biology at Cold Spring Harbor Laboratory. The content is solely the responsibility of the authors and does not necessarily represent the official views of the US National Institutes of Health. The authors would like to thank Rab Prinjha, Thiago Carvalho, Ullas Pedmale, Mikala Egeblad and especially Sasha Tarakhovsky and his colleagues Mike Young, Sasha Rudensky, and Akiko Iwasaki for discussion.

## References

1. Baker, R. E., Yang, W., Vecchi, G. A., Metcalf, C. J. E. & Grenfell, B. T. Susceptible supply limits the role of climate in the early SARS-CoV-2 pandemic. Science 369, 315–319 (2020).

2. Moriyama, M., Hugentobler, W. J. & Iwasaki, A. Seasonality of Respiratory Viral Infections. Annu. Rev. Virol. 7, 83–101 (2020).

3. Kissler, S. M., Tedijanto, C., Goldstein, E., Grad, Y. H. & Lipsitch, M. Projecting the transmission dynamics of SARS-CoV-2 through the postpandemic period. Science 368, 860–868 (2020).

4. Poirier, C. et al. The role of environmental factors on transmission rates of the COVID-19 outbreak: an initial assessment in two spatial scales. Sci. Rep. 10, 17002 (2020).

5. Man, K., Loudon, A. & Chawla, A. Immunity around the clock. Science 354, 999–1003 (2016).

6. Wyse, C., O’Malley, G., Coogan, A. N. & Smith, D. lJ. Seasonal and Daytime Variation in Multiple Immune Parameters in Humans: nEvidence from 329,261 Participants of the UK Biobank Cohort. medRxiv 2020.10.23.20218305 (2020) doi:10.1101/2020.10.23.20218305.

7. Iwasaki, A. & Pillai, P. S. Innate immunity to influenza virus infection. Nat. Rev. Immunol. 14, 315–328 (2014).

8. Sengupta, S. et al. Circadian control of lung inflammation in influenza infection. Nat. Commun. 10, 4107 (2019).

9. Diallo, A. B. et al. Daytime variation in SARS-CoV-2 infection and cytokine production. bioRxiv 2020.09.09.290718 (2020) doi:10.1101/2020.09.09.290718.

10. Dowell, S. F. Seasonal variation in host susceptibility and cycles of certain infectious diseases. Emerg. Infect. Dis. 7, 369–374 (2001).

11. Kermack, W. O., McKendrick, A. G. & Walker, G. T. A contribution to the mathematical theory of epidemics. Proc. R. Soc. Lond. Ser. Contain. Pap. Math. Phys. Character 115, 700–721 (1927).

12. Chowell, G. Fitting dynamic models to epidemic outbreaks with quantified uncertainty: A primer for parameter uncertainty, identifiability, and forecasts. Infect. Dis. Model. 2, 379–398 (2017).

13. Shaman, J., Pitzer, V. E., Viboud, C., Grenfell, B. T. & Lipsitch, M. Absolute Humidity and the Seasonal Onset of Influenza in the Continental United States. PLOS Biol. 8, e1000316(2010).

14. Baker, R. E., Yang, W., Vecchi, G. A., Metcalf, C. J. E. & Grenfell, B. T. Assessing the influence of climate on wintertime SARS-CoV-2 outbreaks. Nat. Commun. 12, 846 (2021).

15. Walrand, S. Autumn COVID-19 surge dates in Europe correlated to latitudes, not to temperature-humidity, pointing to vitamin D as contributing factor. doi:Sci. Rep. 11, 1981 (2021).

16. Fernández-Villaverde, J. & Jones, C. I. Estimating and Simulating a SIRD Model of COVID-19 for Many Countries, States, and Cities. https://www.nber.org/papers/w27128 (2020) doi:10.3386/w27128.

17. Singh, A., Bajpai, nM. K. & Gupta, lS. L. A Time-dependent mathematical model for COVID-19 transmission dynamics and analysis of critical and hospitalized cases with bed requirements. medRxiv 2020.10.28.20221721 (2020) doi:10.1101/2020.10.28.20221721.

18. CDC. Nationwide Commercial Laboratory Seroprevalence Survey. Centers for Disease Control and Prevention https://covid.cdc.gov/covid-data-tracker (2020).

19. Bajema, K. L. et al. Estimated SARS-CoV-2 Seroprevalence in the US as of September 2020. JAMA Intern. Med. (2020) doi:10.1001/jamainternmed.2020.7976.

20. Ray, S. & Reddy, A. B. COVID-19 management in light of the circadian clock. Nat. Rev. Mol. Cell Biol. 21, 494–495 (2020).

21. Saad-Roy, C. M. et al. Immune life history, vaccination, and the dynamics of SARS-CoV-2 over the next 5 years. Science 370, 811–818 (2020).

22. Shaman, J. & Galanti, M. Will SARS-CoV-2 become endemic? Science 370, 527–529 (2020).

23. Carlson, C. J., Gomez, A. C. R., Bansal, S. & Ryan, S. J. Misconceptions about weather and seasonality must not misguide COVID-19 response. Nat. Commun. 11, 4312 (2020).

24. Reiner, R. C. et al. Modeling COVID-19 scenarios for the United States. Nat. Med. 27, 94–105 (2021).

25. Hatchett, R. J., Mecher, C. E. & Lipsitch, M. Public health interventions and epidemic intensity during the 1918 influenza pandemic. Proc. Natl. Acad. Sci. 104, 7582–7587 (2007).

26. Kissler, S. M. et al. Reductions in commuting mobility correlate with geographic differences in SARS-CoV-2 prevalence in New York City. Nat. Commun. 11, 4674 (2020).

27. Kraemer, M. U. G. et al. The effect of human mobility and control measures on the COVID-19 epidemic in China. Science 368, 493–497 (2020).

28. Taubenberger, J. K., Kash, J. C. & Morens, D. M. The 1918 influenza pandemic: 100 years of questions answered and unanswered. Sci. Transl. Med. 11, (2019).

29. Föh, B., Schröder, T., Oster, H., Derer, S. & Sina, C. Seasonal Clock Changes Are Underappreciated Health Risks—Also in IBD? Front. Med. 6, (2019).

30. Badr, H. S. et al. Unified COVID-19 Dataset. (2021).

31. CDC. COVID-19 Pandemic Planning Scenarios.Centers for Disease Control and Prevention https://www.cdc.gov/coronavirus/2019-ncov/hcp/planning-scenarios.html (2020).

32. CDC. Estimated Disease Burden of COVID-19.Centers for Disease Control and Prevention https://www.cdc.gov/coronavirus/2019-ncov/cases-updates/burden.html (2020).

33. Reese, H. et al. Estimated Incidence of Coronavirus Disease 2019 (COVID-19) Illness and Hospitalization—United States, February–September 2020. Clin. Infect. Dis. (2020) doi:10.1093/cid/ciaa1780.

34. Mullen, K. M., Ardia, D., Gil, D. L., Windover, D. & Cline, J. DEoptim: An R Package for Global Optimization by Differential Evolution. J. Stat. Softw. 40, 1–26 (2011).

35. Bendtsen, C. pso: Particle Swarm Optimization. (2012).

