## Supplementary Materials for "Circadian immunity, sunrise time and the seasonality of respiratory infections"

### Data and code availability

All data and code used in this study are publicly available (github.com/ziyimo/circ-immu).

### Data collection

Epidemiological data of influenza at the state level since the 2010-11 season were acquired from FluView, a weekly influenza surveillance report by the CDC (www.cdc.gov/flu/weekly). These include the numbers of total weekly tests and positive influenza A/B tests reported by WHO/NREVSS Collaborating Labs, as well as statistics for influenza-like illness (ILI) visits from the US Outpatient Influenza-like Illness Surveillance Network (ILINet). State-level COVID-19 death and hospitalization data were obtained from The COVID Tracking Project (covidtracking.com/data/download/all-states-history.csv). The number of COVID-19-related deaths in 31 major cities worldwide was acquired from the Unified COVID-19 Dataset[30] and the UK government COVID-19 website (https://api.coronavirus.data.gov.uk). To account for the periodic oscillations in reported deaths, we applied a seven day moving average to the data. In addition, extreme single day outlier values in Madrid, Delhi, Mexico City and Santiago were replaced with local average values from the six closest days.

Day lengths and sunrise times were calculated with the *astral* package (pypi.org/project/astral/), whereas near-surface specific humidity came from the ERA5 dataset (doi: 10.24381/cds.20d54e34). Following ref. [1], day length, sunrise and humidity variables at the state level were calculated as population-weighted averages using population density in 2015 reported in GPWv4 from CIESIN (doi: 10.7927/H49C6VHW). Weighting was not conducted for the city level environmental variables, and in this case day length and sunrise were based on the *geopy* (pypi.org/project/geopy/) longitude and latitude for each city. Our $R_0$ models aim to capture broad patterns of seasonal dynamics and therefore year-to-year variations in the climatological variable were disregarded by averaging over 5 years of humidity data from 2014 to 2018. Since the day length and sunrise variables exhibit negligible year-to-year variation, we used values calculated by *astral* for the year 2019. Curated data used to fit the model are available via github (github.com/ziyimo/circ-immu/tree/main/state_lv_data).

### SIRS model for influenza

We applied a classical SIRS framework[1,11] (**Supplementary Fig. S2A**) to model the epidemiology of influenza as an endemic. The susceptible ($S$), infected ($I$) and recovered ($R$) compartments in the population ($N$) are governed by the following differential equations:

$$\frac{dS}{dt} = \frac{R}{L} - \frac{\beta(t)IS}{N}$$
$$\frac{dI}{dt} = \frac{\beta(t)IS}{N} - \frac{I}{D}$$
$$\frac{dR}{dt} = \frac{I}{D} - \frac{R}{L}$$

We treated $D$, the mean infection period and $L$, the duration of immunity as known for a certain disease (e.g., $D$ = 5 days and $L$ = 40 weeks for flu[1]). Therefore, the transmission dynamics are driven by the time-varying reproductive number $R_0(t)$, which is simply a scaled version of the contact rate $\beta(t)$, $R_0(t) = \beta(t)D$.

### Modified SIRS model for COVID-19

We modeled COVID-19 death counts data following ref. [16]. In addition, an exposed ($E$) compartment was introduced to account for the non-trivial incubation period of COVID-19. The flow of population among the susceptible ($S$), exposed ($E$), infected ($I$), resolving ($G$) and removed ($R_{em}$) compartments are defined by (see also **Supplementary Fig. S2B**):

$$\frac{dS}{dt} = -\frac{\beta(t)IS}{N}$$
$$\frac{dE}{dt} = \frac{\beta(t)IS}{N} - \sigma^{-1}E$$
$$\frac{dI}{dt} = \sigma^{-1}E - \gamma^{-1}I$$
$$\frac{dG}{dt} = \gamma^{-1}I - \psi^{-1}G$$
$$\frac{dR_{em}}{dt} = \psi^{-1}G$$

Here, we introduced an incubation period $\sigma$ = 5 days, a mean infection period (time from symptom onset to patient isolation) $\gamma$ = 5 days and a mean time of case resolution $\psi$ = 10 days[16,31]. As before, the time-varying contact rate—here given by $\beta(t) = R_0(t) / \gamma$—was fitted to data through our $R_0$ models. The number of newly reported deaths on a particular day was treated as a binomial sample from the *G* compartment on that day with a probability equal to the resolving death rate $\alpha = \delta\psi^{-1}$ = 0.001 (see discussions of model likelihood below for details). This corresponds to a death rate of 1% (we also fitted the models under alternative death rates of 0.7% and 0.5%, see **Supplementary Tables S3 & S4**)[16].

A simplified version of the model proposed in ref.[17] was used to model hospitalizations (**Supplementary Fig. S2C**), as follows:

$$\frac{dS}{dt} = -\frac{\beta(t)IS}{N}$$
$$\frac{dE}{dt} = \frac{\beta(t)IS}{N} - \sigma^{-1}E$$
$$\frac{dI}{dt} = \sigma^{-1}E - \eta\gamma^{-1}I - (1-\eta)\gamma^{-1}I$$
$$\frac{dH}{dt} = \eta\gamma^{-1}I - \kappa^{-1}H$$
$$\frac{dR_{em}}{dt} = (1-\eta)\gamma^{-1}I + \kappa^{-1}H$$

with $\sigma$ and $\gamma$ defined the same way as above, and additional parameters including a hospitalization rate $\eta$ = 2.6% (we also fitted the models under an alternative value of 5%, see **Supplementary Tables S5-S7; Supplementary Figs. S4-S6**) and a mean length of hospitalization $\kappa$ = 10 days[31–33].

### Models for $R_0(t)$

The compartmental models allowed us to model the epidemiological dynamics as driven by some choice of underlying variables that directly determine $R_0(t)$. We could subsequently evaluate the goodness of fit by calculating the model likelihood. For the model comparison study, we considered the following $R_0$ models.

1. **A sinusoidal $R_0$ model (base)**

$$R_0(t) = \cos\left(\frac{2\pi}{364}(t-\phi)\right)\frac{(R_{0,max} - R_{0,min})}{2} + \frac{(R_{0,max} + R_{0,min})}{2}$$

In this model, $R_0$ depends solely on the day of the year $t$, and fluctuates annually between $R_{0,min}$ and $R_{0,max}$ in a sinusoidal wave. A single free parameter $\phi \in [0,\ 364)$ which governs the day of the year when $R_0$ reaches its maximum is fitted to the data. This was considered the base model in the model comparison study.

2. **A climate $R_0$ model (h)**

$$R_0(t) = R_{0,min} + e^{\alpha_1 h(t)} R_{0,range}$$

This is the climate model from refs. [1,13]. The climate dependency parameter $\alpha_1 < 0$ governs the rate of exponential decay in $R_0$ as the specific humidity $h$ increases. We also experimented with a Gaussian form for the climate model and found that the model in its original exponential form provided a better fit.

3. **A day-length $R_0$ model (d)**

$$R_0(t) = R_{0,min} + e^{\alpha_2 (d(t) - d_0)^2} R_{0,range}$$

The Gaussian day-length model estimates $d_0 \in [0,\ 1]$, the value of day length when $R_0$ attains its maximum, from the data, and therefore permits either a monotonic or peaked relationship between $R_0$ and the day length $d$. The width of the curve is governed by $\alpha_2 < 0$. In practice, the best-fit models specified a monotonically decreasing relationship between $R_0$ and day length ($d_0 < d_{min}$, where $d_{min}$ is the minimum observed value of the day length covariate, see **Supplementary Tables S1, S2 & S5**).

4. **A sunrise/day-length $R_0$ model (sd)**

$$R_0(t) = R_{0,min} + e^{\alpha_3(s(t)-s_0)^2+\alpha_2(d(t)-d_0)^2} R_{0,range}$$

A composite model with $\alpha_3 < 0$ controlling the dependence of $R_0$ on sunrise time *s*, and $\alpha_2 < 0$ controlling the dependence of $R_0$ on day length *d*. $s_0$ is defined in a similar fashion to $d_0$ described above. However, in contrast to $d_0$, best fit values of $s_0$ did specify “peaks” of $R_0$ (**Supplementary Tables S1, S4 & S7**).

5. **A climate/day-length $R_0$ model (hd)**

$$R_0(t) = R_{0,min} + e^{\alpha_1 h(t)+\alpha_2(d(t)-d_0)^2} R_{0,range}$$

A composite model with $\alpha_1 < 0$ controlling the dependence of $R_0$ on specific humidity *h*, and $\alpha_2 < 0$ controlling the dependence of $R_0$ on day length *d*.

6. **A climate/sunrise/day-length $R_0$ model (hsd)**

$$R_0(t) = R_{0,min} + e^{\alpha_1 h(t)+\alpha_3(s(t)-s_0)^2+\alpha_2(d(t)-d_0)^2} R_{0,range}$$

The most comprehensive composite model with $\alpha_1 < 0$ controlling the dependence of $R_0$ on specific humidity *h*, $\alpha_3 < 0$ controlling the dependence of $R_0$ on sunrise time *s, and* $\alpha_2 < 0$ controlling the dependence of $R_0$ on day length *d*.

For all of the above models, the day length *d* was scaled between 0 (0 hr) and 1 (24 hrs) whereas sunrise time *s* was scaled between 0 (midnight) and 1 (noon). For influenza models, $R_{0,max}$ was fixed to 2 and $R_{0,min}$ was fixed to 1.2, based on reproductive numbers from the literature and an expected largest reduction in $R_0$ of 40%[1]. The range of $R_0$ values for COVID, on the other hand, was fitted to data.

*Model likelihood for influenza testing data*

The epidemiological data for flu consist of the total number of tests conducted in a week, $N_t$, the number of positive tests, $k_t$, and the proportion of Influenza-like illness (ILI) visits, $\pi_t$. We modeled testing as a binomial sampling process, such that $k_t \sim B(N_t,\, q_t)$, where $q_t$ is the probability of testing positive in week *t*. We further assume that $q_t$ is proportional to $p_t$, the rate of flu infection in the general population in week *t,* which can be obtained from our model as the proportion of the population that falls in the *I* compartment in week t (we use the daily value corresponding to day 4 of week *t*). Indeed, we could define $q_t = \frac{p_t}{\pi_t}$ if it were true that 1) the rate of flu infection among hospital visitors equals that in the general population, and 2) all true positive flu cases exhibit symptoms and register as ILI visits. For various reasons, however, we expect these assumptions to be too restrictive and to unrealistically inflate $q_t$, so we allowed for a free parameter $c \in (0,\, 1]$ to serve as a flexible constant of proportionality to be estimated from the data, that is, $q_t = \frac{c \times p_t}{\pi_t}$ .

A complication with this strategy is that, unless *c* is strictly bounded, some choices of model parameters still may lead to $q \geq 1$, making the binomial likelihood ill-defined. Furthermore, strictly capping *c* so that this scenario never occurs is too restrictive, owing to noise in the data and the need to explore a broad range of parameter values during optimization. To address this problem, we adopted a penalized likelihood strategy for truncation of *q,* forcing it to remain below a designated cap $q_{cap}$ but penalizing the truncation when it is applied. Specifically, we assume a penalized negative log likelihood as follows:

$$
\begin{aligned}
-\sum_t \log \mathcal{L}_{pnl}(q_t; k_t, N_t) &= -\sum_t \log \mathcal{L}(q'_t; k_t, N_t) + \sum_t \lambda(q_t - q'_t) \\
&= -\sum_t \log \Pr(k_t | q'_t, N_t) + \sum_t \lambda(q_t - q'_t)
\end{aligned}
$$

where $q'_t = \min(q_t, q_{cap})$ and the hyperparameter $\lambda$ determines how heavily the model is penalized for exceeding the cap. $q_{cap}$ was set to either the upper 99.9% binomial confidence interval calculated from the maximum test positivity rate in the data, or 0.999, whichever is lower. As noted in the text, we experimented with various values of $\lambda$ and found that our results were fairly insensitive to this parameter (we focus on the case of $\lambda$=10 in our main results).

For the purpose of plotting the raw data (as in **Fig. 2**), we used an estimator for the rate of flu infection in the general population in week *t* given by $\hat{p_t} = \frac{1}{c}\frac{k_t}{N_t}\pi_t$, where *c* was estimated in our penalized likelihood framework In addition, we used this same estimator $\hat{p}_t$ to calculate correlation coefficients between the raw data and our climate model (h) fit,to allow comparison with a previous study[13]. We found that modeling the specific humidity covariate alone, our model achieved a similar Pearson's correlation to the data of about *r* = 0.7.

*Model likelihoods for COVID-19 death and hospitalization data*

The difference in the nature of death and hospitalization data necessitated slightly different approaches to calculating model likelihood. Death data consists of the daily number of *newly reported* fatalities. We could therefore model the absolute number of deaths on each day as a binomial sample of the resolving cases on that day. This results in a very similar likelihood model to that of the flu testing data. Formally, using a probability of $a = \delta\psi^{-1}$ (resolving death rate) and a sample size of $G_t$ (number of resolving cases on day *t* predicted by the model), the probability of observing $d_t$ deaths follows a $B(G_t,\, \alpha)$ distribution. To accommodate likelihood calculation of extremely misspecified models proposed during the optimization process, we used an adjusted sample size $G_t' = d_t$ in cases where $G_t < d_t$, and explicitly penalize such cases in our likelihood calculation as follows:

$$-\sum_t \log \mathcal{L}(G_t; d_t) = -\sum_t \log \Pr\big(d_t|\alpha, G'_t\big) + \sum_t \big(G'_t - G_t\big)$$

where $G'_t = \max(G_t, d_t)$. The hospitalization counts, on the other hand, are cumulative measures that reflect the *current* number of hospitalized patients and therefore were modeled by the *H* compartment itself. We therefore treated the number of current hospitalization as a homoscedastic observation of the $H$ compartment, and calculated model likelihood using a normal probability distribution with uniform variance.

*Model fitting and evaluation*

An instance of the flu SIRS model was run for 50 years, all states in parallel, to allow it to reach an equilibrium. Each state was initialized with a single infected individual in a population corresponding to the 2010 census population of that state. The model was subsequently run with annual $R_0$ values calculated given the set of $R_0$ model parameters (shared among all states) and state-specific variables (humidity, day length, sunrise). In other words, each instance of the model consists of a unified $R_0$ model shared by all states, and a separate compartmental model for each state. For model fitting, we took model output from the last number of years corresponding to the number of years for which data were available.

The COVID-19 hospitalization model (SEIHRem) and death model (SEIGRem) were run from March 13 2020, the day when the COVID-19 pandemic was declared a national emergency in the U.S., until Jan 31 2021. In addition to the $R_0$ model parameters, the initial proportion of infected individuals for each state or city ($I_{init}/N$) were treated as free in these models. In the case of $R_0$ models for COVID-19, each of the nine U.S. census divisions was given its own sunrise "peak" $s_0$, resulting in nine separate $s_0$ parameters in total. For the city-level modelling, each city was given its own $s_0$ . The SEIHRem and SEIGRem models were run jointly for multiple geographic units in an otherwise similar fashion to the SIRS model.

Optimization was conducted using the "DEoptim"[34] and "pso"[35] packages in R, which implement the differential evolution and particle swarm optimization algorithms, respectively. We fitted the 6 distinct $R_0$ models to flu surveillance data in 40 states with fewer than 200 weeks of missing data (out of 520 weeks total from the 2010-11 season to the 2019-20 season), COVID-19 hospitalization data in 48 contiguous states (plus Hawaii and DC), and COVID-19 death data in 46 contiguous states (plus DC) until Jan 31 2021. States with fewer than 700 total deaths (AK, HI, VT, WY) were excluded from COVID-19 death data. Finally, model fit was evaluated using Bayesian Information Criterion $\mathrm{BIC} = k\ln(n) - 2\ln(\widehat{L})$, where $k$ is the number of model parameters, $n$ is the number of data points and $\hat{L}$ is the maximized likelihood of the model.

*Forward simulations*

To estimate the impact of daylight saving time changes on the endemic transmission dynamics of flu, we ran forward simulations of the SIRS model with best-fit parameters $\hat{\theta}^{\mathbf{ML}}$ of the sunrise/day-length (sd) $R_0$ model, estimated from 10 years worth of data. The model, with its parameters set at the MLE, was run for 50 years using annual cycles of the covariates as described previously. Finally, model prediction for the last year (which reflects the annual pattern under an endemic steady-state) was taken to demonstrate the dynamics of a typical flu season. We considered three alternative scenarios for sunrise time where 1) states follow their current DST schedule (with AZ and HI opting out), 2) all states implement permanent standard time, and 3) all states implement permanent DST.

We gauged the uncertainty in the counterfactual model predictions in two ways. First, replicates of each set of simulations were performed with model parameters resampled from a multivariate Gaussian distribution $\theta \sim \mathcal{N}\left(\hat{\theta}^{\mathbf{ML}}, \mathbf{\Sigma}\right)$, where the covariance matrix is estimated as the inverse of the negative Hessian of the log-likelihood function at $\hat{\theta}^{ML}$, $\mathbf{\Sigma} = \left[-\mathbf{H}\log\mathcal{L}\left(\hat{\theta}^{\mathbf{ML}}\right)\right]^{-\mathbf{1}}$. The Hessian was calculated numerically by finite difference approximation with the *optimHess* function in R. Second, we also performed replicates of simulations with bootstrapped MLE model parameters[12], which were obtained by optimizations under datasets consisting of jurisdictions (i.e. states, or cities) resampled with replacement. The standard deviation of bootstrap samples was used to report the variance in model prediction, unless specified otherwise. Similarly, simulations under these two approaches were performed to estimate the counterfactual effect of time change on COVID epidemiology.

It is worth noting that estimates of parameter variance from the Hessian are based solely on how sharply the likelihood surface is peaked around the MLE, and therefore cannot account for uncertainty due to multi-modality of the likelihood surface, which is very likely the case for our covariate-dependent $R_0$ models. The bootstrapping procedure outlined above fares better at estimating the true uncertainty in the model predictions. However, as discussed in the text, neither approach is able to address the additional uncertainty due to modeling assumptions[12].

**Supplementary Table S1. Maximum likelihood, best-fit parameters and BIC of $R_0$ models fitted to U.S. influenza surveillance data**

| **λ=10** | | | | | |
|---|---|---|---|---|---|
| **Model** | **-log(*L*)** | **c** | **$R_0$ model parameters** | ***k*** | **BIC** |
| base | 1166629.845 | 0.314597 | $\varphi$ = 336.091444 | 2 | 2333279.587 |
| h | 1089364.253 | 0.305390 | $\alpha$ = -76.063888 | 2 | 2178748.403 |
| d | 728915.912 | 0.338684 | $\alpha$ = -4.284229, $d_0$ = 0.000077 | 3 | 1457861.669 |
| sd | 708852.803 | 0.373150 | $\alpha_1$ = -20.014690, $s_0$ = 0.534814, $\alpha_2$ = -5.932632, $d_0$ = 0.093795 | 5 | 1417755.348 |
| hd | 728997.776 | 0.338188 | $\alpha_1$ = -0.046645, $\alpha_2$ = -4.278007, $d_0$ = 0.001025 | 4 | 1458035.347 |
| hsd | 706595.238 | 0.353127 | $\alpha_1$ = -12.889948, $\alpha_2$ = -18.820708, $s_0$ = 0.537953, $\alpha_3$ = -4.296793, $d_0$ = 0.063486 | 6 | 1413250.167 |
| **λ=1000** | | | | | |
| **Model** | **-log(*L*)** | **c** | **$R_0$ model parameters** | ***k*** | **BIC** |
| base | 1530948.860 | 0.183754 | $\varphi$ = 337.589711 | 2 | 3061917.617 |
| h | 1534727.097 | 0.160003 | $\alpha$ = -75.613082 | 2 | 3069474.091 |
| d | 1011341.089 | 0.242170 | $\alpha$ = -4.268851, $d_0$ = 0.000744 | 3 | 2022712.024 |
| sd | 1000324.223 | 0.240695 | $\alpha_1$ = -17.920377, $s_0$ = 0.549864, $\alpha_2$ = -5.063578, $d_0$ = 0.060337 | 5 | 2000698.187 |
| hd | 1011439.152 | 0.242286 | $\alpha_1$ = -0.057132, $\alpha_2$ = -4.265932, $d_0$ = 0.000179 | 4 | 2022918.098 |
| hsd | 1000520.782 | 0.239310 | $\alpha_1$ = -3.356418, $\alpha_2$ = -18.635970, $s_0$ = 0.551916, $\alpha_3$ = -4.715343, $d_0$ = 0.053300 | 6 | 2001101.255 |

**Supplementary Table S2. Maximum likelihood, select best-fit parameters* and BIC of $R_0$ models fitted to U.S. state-level COVID-19 death data under a death rate ($\delta$) of 1%.**

| Model | -log(*L*) | $R_{0, min}$ | $R_{0, range}$ | $R_0$ model parameters | *k* | BIC |
|---|---|---|---|---|---|---|
| base | 590485.8 | 0.800 | 1.2 | $\varphi$ = 348.5186 | 50 | 1181180.732 |
| h | 213127.0 | 1.004 | 1.2 | $\alpha$ = -300 | 50 | 426463.1321 |
| d | 117021.1 | 0.802 | 1.202 | $\alpha$ = -5.041, $d_0$ = 0 | 51 | 234255.5148 |
| sd | 103736.5 | 0.837 | 0.600 | $\alpha_1$ = -35.185, $s_0$†, $\alpha_2$ = -19.656, $d_0$ = 0.351 | 61 | 207728.1412 |
| hd | 117066.4 | 0.800 | 1.2 | $\alpha_1$ = -0.007060576, $\alpha_2$ = -5.014881, $d_0$ = 8.727869e-05 | 52 | 234350.2974 |
| hsd | 103990.2 | 0.870 | 0.601 | $\alpha_1$ = -0.00101663, $\alpha_2$ = -14.80779, $s_0$†, $\alpha_3$ = -42.8548, $d_0$ = 0.4111704 | 62 | 208239.7239 |

* see **Supplementary Table S3** for $I_{init}/N$ values of the sd model
†see **Supplementary Table S4** for $s_0$ values of the sd model

**Supplementary Table S3. COVID-19 death sd model fit of the initial proportion of infected population ($I_{init}/N$)**

| State | Death rate (*δ*) | | | State | Death rate (*δ*) | | |
|---|---|---|---|---|---|---|---|
| | 1% | 0.7% | 0.5% | | 1% | 0.7% | 0.5% |
| AL | 9.51E-04 | 2.38E-03 | 2.84E-03 | NC | 1.07E-03 | 2.07E-03 | 3.15E-03 |
| AR | 1.33E-03 | 1.71E-03 | 1.80E-03 | ND | 2.65E-03 | 2.40E-03 | 2.30E-03 |
| AZ | 2.11E-03 | 3.07E-03 | 3.31E-03 | NE | 1.11E-03 | 1.34E-03 | 1.53E-03 |
| CA | 1.03E-03 | 1.63E-03 | 2.02E-03 | NH | 6.53E-04 | 7.39E-03 | 9.54E-03 |
| CO | 1.33E-03 | 3.31E-03 | 4.42E-03 | NJ | 1.10E-02 | 1.22E-02 | 1.33E-02 |
| CT | 7.94E-03 | 9.76E-03 | 1.06E-02 | NM | 1.13E-03 | 1.87E-03 | 2.28E-03 |
| DC | 6.43E-03 | 8.90E-03 | 9.67E-03 | NV | 1.79E-03 | 3.33E-03 | 3.89E-03 |
| DE | 3.58E-03 | 5.68E-03 | 6.83E-03 | NY | 1.04E-02 | 1.29E-02 | 1.37E-02 |
| FL | 2.33E-03 | 3.28E-03 | 3.45E-03 | OH | 1.43E-03 | 2.25E-03 | 3.80E-03 |
| GA | 2.63E-03 | 4.10E-03 | 4.57E-03 | OK | 8.80E-04 | 1.10E-03 | 1.22E-03 |
| IA | 1.91E-03 | 2.14E-03 | 2.36E-03 | OR | 9.77E-04 | 1.25E-03 | 1.16E-03 |
| ID | 1.63E-03 | 1.80E-03 | 1.51E-03 | PA | 3.37E-03 | 5.44E-03 | 5.66E-03 |
| IL | 3.76E-03 | 4.64E-03 | 5.35E-03 | RI | 5.41E-03 | 6.88E-03 | 7.46E-03 |
| IN | 2.45E-03 | 3.43E-03 | 3.63E-03 | SC | 2.25E-03 | 3.39E-03 | 3.70E-03 |
| KS | 1.01E-03 | 1.13E-03 | 1.25E-03 | SD | 2.00E-03 | 2.09E-03 | 1.59E-03 |
| KY | 9.99E-04 | 1.51E-03 | 1.38E-03 | TN | 1.17E-03 | 1.64E-03 | 1.64E-03 |
| LA | 4.99E-03 | 6.44E-03 | 6.48E-03 | TX | 1.63E-03 | 2.40E-03 | 2.62E-03 |
| MA | 8.31E-03 | 9.20E-03 | 9.83E-03 | UT | 6.51E-04 | 7.25E-04 | 5.58E-04 |
| MD | 3.46E-03 | 5.75E-03 | 5.57E-03 | VA | 9.17E-04 | 2.70E-03 | 4.91E-03 |
| ME | 2.47E-04 | 6.28E-03 | 9.23E-03 | WA | 1.71E-03 | 2.15E-03 | 2.32E-03 |
| MI | 3.45E-03 | 4.09E-03 | 4.41E-03 | WI | 1.25E-03 | 1.40E-03 | 2.96E-03 |
| MN | 2.05E-03 | 2.35E-03 | 2.61E-03 | WV | 1.12E-03 | 1.16E-03 | 1.07E-03 |
| MO | 9.41E-04 | 2.57E-03 | 3.46E-03 | | | | |
| MS | 2.92E-03 | 3.78E-03 | 3.87E-03 | | | | |
| MT | 1.83E-03 | 2.14E-03 | 1.35E-03 | | | | |

**Supplementary Table S4. COVID-19 death sd model fit of the peak of sunrise effect ($s_0$)**

| Census division | Death rate ($\delta$) | | |
|---|---|---|---|
| | **1%** | **0.7%** | **0.5%** |
| Pacific | 0.4357 | 0.4376 | 0.4468 |
| East South Central | 0.4659 | 0.4700 | 0.4792 |
| West South Central | 0.4698 | 0.4815 | 0.4891 |
| Mountain | 0.4660 | 0.4779 | 0.4998 |
| New England | 0.6142 | 0.5959 | 0.5667 |
| South Atlantic | 0.4542 | 0.4676 | 0.4837 |
| West North Central | 0.4940 | 0.4926 | 0.5110 |
| East North Central | 0.4894 | 0.4946 | 0.5194 |
| Middle Atlantic | 0.6543 | 0.6222 | 0.6016 |

**Supplementary Table S5. Maximum likelihood, select best-fit parameters* and BIC of $R_0$ models fitted to U.S. state-level COVID-19 hospitalization data**

| η=0.026 | | | | | | |
|---|---|---|---|---|---|---|
| **Model** | **-log(*L*)** | $R_{0,\ min}$ | $R_{0,\ range}$ | **$R_0$ model parameters** | ***k*** | **BIC** |
| base | 7434203253 | 0.9475 | 0.4381 | φ = 324.4393 | 53 | 1.4868E+10 |
| h | 15607111315 | 0.7931 | 0.4472 | α = -15.20934 | 53 | 3.1214E+10 |
| d | 6866896580 | 0.9755 | 0.4397 | α = -99.63272, $d_0$ = 0.4344352 | 54 | 1.3734E+10 |
| sd | 5798075108 | 0.9998 | 0.6812 | $α_1$ = -35.16802, $s_0$†, $α_2$ = -147.2332, $d_0$ = 0.4378637 | 64 | 1.1596E+10 |
| hd | 6848815355 | 0.9900 | 0.4236 | $α_1$ = -0.07583849, $α_2$ = -105.771, $d_0$ = 0.4332639 | 55 | 1.3698E+10 |
| hsd | 5854646819 | 1.0401 | 0.7870 | $α_1$ = -0.06285446, $α_2$ = -49.1765, $s_0$†, $α_3$ = -283.1534, $d_0$ = 0.4364146 | 65 | 1.1709E+10 |
| **η=0.05** | | | | | | |
| **Model** | **-log(*L*)** | $R_{0,\ min}$ | $R_{0,\ range}$ | **$R_0$ model parameters** | ***k*** | **BIC** |
| base | 6793366617 | 0.9265 | 0.3346 | φ = 315.5494 | 53 | 1.3587E+10 |
| h | 12266896845 | 0.9141 | 0.3277 | α = -39.19565 | 53 | 2.4534E+10 |
| d | 6646509850 | 0.9535 | 0.3321 | α = -106.2214, $d_0$ = 0.4413445 | 54 | 1.3293E+10 |
| sd | 6083071230 | 0.9877 | 0.5050 | $α_1$ = -18.58159, $s_0$†, $α_2$ = -332.4335, $d_0$ = 0.4488209 | 64 | 1.2166E+10 |
| hd | 6533551013 | 0.9837 | 0.3367 | $α_1$ = -0.1135141, $α_2$ = -190.3919, $d_0$ = 0.4428364 | 55 | 1.3067E+10 |
| hsd | 6082344989 | 0.9917 | 0.5267 | $α_1$ = -0.01785445, $α_2$ = -9.442807, $s_0$†, $α_3$ = -370.0165, $d_0$ = 0.4491905 | 65 | 1.2165E+10 |

* see **Supplementary Table S6** for $I_{init}/N$ values
†see **Supplementary Table S7** for $s_0$ values of sd and hsd models

**Supplementary Table S6. COVID-19 hospitalization model fit of the initial proportion of infected population ($I_{init}/N$)**

| | η=0.026 | | | | | | η=0.05 | | | | | |
|---|---|---|---|---|---|---|---|---|---|---|---|---|
| | base | h | d | sd | hd | hsd | base | h | d | sd | hd | hsd |
| AL | 6.29E-03 | 7.86E-04 | 3.23E-03 | 4.76E-03 | 3.47E-03 | 6.08E-03 | 4.04E-03 | 1.07E-03 | 2.32E-03 | 3.91E-03 | 2.77E-03 | 3.45E-03 |
| AZ | 5.14E-03 | 2.94E-04 | 2.72E-03 | 3.58E-03 | 2.64E-03 | 4.23E-03 | 4.38E-03 | 3.09E-04 | 2.60E-03 | 3.61E-03 | 3.16E-03 | 3.71E-03 |
| AR | 7.20E-03 | 5.96E-04 | 3.45E-03 | 4.37E-03 | 3.45E-03 | 5.03E-03 | 2.65E-03 | 6.40E-04 | 1.49E-03 | 1.93E-03 | 1.74E-03 | 2.01E-03 |
| CA | 6.32E-03 | 2.39E-04 | 3.57E-03 | 3.45E-03 | 3.45E-03 | 4.57E-03 | 3.17E-03 | 2.51E-04 | 1.98E-03 | 2.46E-03 | 2.50E-03 | 2.54E-03 |
| CO | 2.19E-04 | 8.38E-05 | 5.21E-03 | 5.31E-03 | 5.26E-03 | 5.90E-03 | 8.74E-04 | 5.07E-05 | 9.26E-04 | 1.09E-03 | 2.14E-03 | 1.31E-03 |
| CT | 1.27E-02 | 1.76E-04 | 6.18E-03 | 9.09E-03 | 6.42E-03 | 9.40E-03 | 8.34E-03 | 1.69E-04 | 3.86E-03 | 4.28E-03 | 4.13E-03 | 4.13E-03 |
| DE | 2.52E-03 | 2.68E-04 | 3.99E-03 | 4.65E-03 | 3.98E-03 | 5.00E-03 | 5.80E-03 | 2.99E-04 | 2.69E-03 | 2.79E-03 | 2.88E-03 | 2.71E-03 |
| DC | 7.16E-02 | 1.44E-02 | 7.90E-03 | 8.47E-03 | 7.93E-03 | 9.03E-03 | 1.15E-02 | 1.51E-03 | 6.49E-03 | 8.01E-03 | 7.60E-03 | 8.59E-03 |
| FL | 1.14E-02 | 1.87E-03 | 5.76E-03 | 7.29E-03 | 5.81E-03 | 8.97E-03 | 1.15E-02 | 2.11E-03 | 6.81E-03 | 9.71E-03 | 7.54E-03 | 9.68E-03 |
| GA | 6.93E-03 | 7.39E-04 | 3.61E-03 | 3.07E-03 | 3.51E-03 | 3.44E-03 | 4.85E-03 | 9.36E-04 | 2.57E-03 | 3.27E-03 | 3.14E-03 | 3.38E-03 |
| HI | 6.27E-02 | 1.15E-05 | 1.85E-02 | 1.91E-02 | 1.83E-02 | 1.76E-02 | 6.92E-02 | 1.00E-01 | 5.76E-02 | 2.95E-02 | 5.02E-02 | 2.92E-02 |
| ID | 9.51E-03 | 7.18E-05 | 3.34E-03 | 1.91E-03 | 3.53E-03 | 2.14E-03 | 3.85E-04 | 4.47E-05 | 1.33E-03 | 1.67E-03 | 1.39E-03 | 1.61E-03 |
| IL | 1.08E-02 | 7.02E-04 | 5.75E-03 | 6.52E-03 | 5.45E-03 | 8.03E-03 | 8.47E-03 | 5.84E-04 | 4.29E-03 | 5.19E-03 | 4.35E-03 | 5.10E-03 |
| IN | 7.20E-03 | 5.45E-04 | 3.69E-03 | 3.71E-03 | 3.39E-03 | 4.07E-03 | 5.77E-03 | 5.65E-04 | 2.78E-03 | 3.45E-03 | 3.48E-03 | 3.31E-03 |
| IA | 9.33E-03 | 5.02E-04 | 4.56E-03 | 3.18E-03 | 4.55E-03 | 3.46E-03 | 1.10E-03 | 1.26E-04 | 1.71E-03 | 1.52E-03 | 1.83E-03 | 2.00E-03 |
| KS | 1.27E-02 | 3.15E-04 | 6.08E-03 | 3.75E-03 | 5.96E-03 | 4.18E-03 | 1.20E-02 | 3.94E-02 | 5.36E-03 | 8.11E-03 | 5.84E-03 | 6.90E-03 |
| KY | 7.82E-03 | 4.81E-04 | 3.74E-03 | 2.18E-03 | 3.77E-03 | 2.40E-03 | 4.07E-03 | 4.35E-04 | 1.94E-03 | 2.53E-03 | 2.27E-03 | 2.52E-03 |
| LA | 1.07E-02 | 2.03E-03 | 5.55E-03 | 7.17E-03 | 5.60E-03 | 9.32E-03 | 8.67E-03 | 1.89E-03 | 4.30E-03 | 6.52E-03 | 5.20E-03 | 6.26E-03 |
| ME | 1.96E-04 | 2.98E-05 | 5.29E-03 | 8.79E-03 | 5.03E-03 | 8.95E-03 | 4.54E-04 | 3.28E-05 | 3.76E-04 | 4.41E-04 | 3.86E-04 | 3.84E-04 |
| MD | 1.10E-02 | 1.98E-04 | 5.58E-03 | 6.06E-03 | 5.55E-03 | 5.55E-03 | 9.28E-03 | 1.92E-04 | 2.92E-03 | 3.45E-03 | 3.59E-03 | 3.22E-03 |
| MA | 1.58E-02 | 1.43E-04 | 8.36E-03 | 1.03E-02 | 8.27E-03 | 1.17E-02 | 1.19E-02 | 1.33E-04 | 5.75E-03 | 6.57E-03 | 6.19E-03 | 6.69E-03 |
| MI | 1.12E-02 | 2.89E-04 | 4.91E-03 | 3.09E-03 | 4.65E-03 | 3.06E-03 | 7.10E-03 | 1.45E-04 | 2.80E-03 | 2.68E-03 | 2.82E-03 | 2.74E-03 |
| MN | 9.94E-03 | 7.62E-05 | 4.05E-03 | 2.14E-03 | 4.13E-03 | 2.13E-03 | 6.84E-04 | 6.09E-05 | 1.39E-03 | 1.19E-03 | 1.30E-03 | 1.37E-03 |
| MS | 7.48E-03 | 1.67E-03 | 4.46E-03 | 5.70E-03 | 4.41E-03 | 7.15E-03 | 7.01E-03 | 1.84E-03 | 3.86E-03 | 5.88E-03 | 4.57E-03 | 5.84E-03 |
| MO | 6.64E-03 | 6.18E-04 | 3.42E-03 | 3.27E-03 | 3.50E-03 | 3.90E-03 | 5.04E-03 | 7.02E-04 | 2.43E-03 | 3.00E-03 | 2.77E-03 | 3.00E-03 |
| MT | 1.02E-02 | 4.79E-04 | 2.78E-03 | 2.17E-03 | 2.75E-03 | 2.17E-03 | 6.72E-03 | 7.69E-05 | 1.89E-03 | 2.05E-03 | 1.90E-03 | 2.24E-03 |
| NE | 1.00E-02 | 1.00E-01 | 1.00E-01 | 1.00E-01 | 1.00E-01 | 1.00E-01 | 7.11E-03 | 1.00E-01 | 1.00E-01 | 1.00E-01 | 1.00E-01 | 1.00E-01 |
| NV | 4.97E-03 | 3.22E-04 | 2.60E-03 | 4.09E-03 | 2.77E-03 | 5.58E-03 | 6.15E-03 | 2.34E-04 | 2.81E-03 | 3.28E-03 | 3.29E-03 | 3.38E-03 |
| NH | 7.71E-05 | 4.56E-05 | 5.38E-03 | 8.27E-03 | 5.32E-03 | 9.03E-03 | 7.87E-04 | 4.58E-05 | 5.52E-04 | 6.64E-04 | 5.75E-04 | 5.88E-04 |
| NJ | 1.65E-02 | 1.53E-02 | 8.81E-03 | 1.02E-02 | 9.77E-03 | 1.29E-02 | 1.22E-02 | 3.43E-04 | 6.79E-03 | 7.91E-03 | 7.45E-03 | 8.14E-03 |

| | | | | | | | | | | | | |
|---|---|---|---|---|---|---|---|---|---|---|---|---|
| NM | 7.49E-03 | 1.62E-04 | 3.92E-03 | 4.85E-03 | 3.45E-03 | 6.04E-03 | 1.67E-03 | 9.15E-05 | 8.83E-04 | 3.32E-03 | 1.21E-03 | 6.21E-04 |
| NY | 1.86E-02 | 1.64E-02 | 1.04E-02 | 1.08E-02 | 1.08E-02 | 1.35E-02 | 1.24E-02 | 1.85E-04 | 6.51E-03 | 7.90E-03 | 7.34E-03 | 8.15E-03 |
| NC | 3.86E-04 | 2.98E-04 | 4.07E-03 | 4.61E-03 | 4.30E-03 | 4.89E-03 | 1.52E-03 | 3.36E-04 | 7.88E-04 | 1.10E-03 | 8.04E-04 | 1.05E-03 |
| ND | 4.26E-03 | 6.19E-04 | 3.18E-03 | 2.58E-03 | 3.32E-03 | 2.73E-03 | 1.61E-03 | 9.50E-05 | 2.02E-03 | 2.17E-03 | 2.15E-03 | 2.10E-03 |
| OH | 8.01E-03 | 2.94E-04 | 3.54E-03 | 2.56E-03 | 3.61E-03 | 2.36E-03 | 2.52E-03 | 2.28E-04 | 1.85E-03 | 1.82E-03 | 2.14E-03 | 1.70E-03 |
| OK | 6.10E-03 | 5.65E-04 | 3.29E-03 | 2.65E-03 | 3.13E-03 | 2.61E-03 | 2.95E-03 | 6.87E-04 | 1.83E-03 | 2.54E-03 | 2.11E-03 | 2.26E-03 |
| OR | 1.06E-04 | 3.09E-05 | 4.99E-03 | 1.06E-03 | 5.37E-03 | 8.35E-04 | 3.82E-04 | 3.27E-05 | 5.12E-04 | 9.94E-04 | 5.42E-04 | 9.25E-04 |
| PA | 7.94E-03 | 2.29E-04 | 3.63E-03 | 7.38E-03 | 3.56E-03 | 8.93E-03 | 3.57E-03 | 2.07E-04 | 2.07E-03 | 1.20E-03 | 2.14E-03 | 1.31E-03 |
| RI | 8.77E-03 | 2.84E-04 | 4.11E-03 | 7.01E-03 | 4.28E-03 | 7.46E-03 | 3.03E-03 | 2.21E-04 | 2.74E-03 | 2.79E-03 | 3.07E-03 | 2.87E-03 |
| SC | 9.65E-03 | 8.24E-04 | 4.64E-03 | 4.62E-03 | 4.46E-03 | 5.05E-03 | 8.19E-03 | 8.88E-04 | 1.00E-01 | 4.57E-03 | 4.27E-03 | 4.58E-03 |
| SD | 5.60E-03 | 1.00E-01 | 3.34E-03 | 2.57E-03 | 3.04E-03 | 2.76E-03 | 1.02E-02 | 6.64E-04 | 2.67E-03 | 2.84E-03 | 2.77E-03 | 3.22E-03 |
| TN | 6.50E-03 | 6.05E-04 | 3.39E-03 | 3.45E-03 | 3.43E-03 | 3.45E-03 | 3.95E-03 | 7.20E-04 | 2.13E-03 | 2.95E-03 | 2.57E-03 | 2.78E-03 |
| TX | 6.18E-03 | 8.75E-04 | 3.48E-03 | 3.84E-03 | 3.42E-03 | 4.35E-03 | 4.02E-03 | 1.13E-03 | 2.45E-03 | 3.13E-03 | 3.06E-03 | 3.22E-03 |
| UT | 1.41E-02 | 5.45E-05 | 5.54E-03 | 2.65E-03 | 5.73E-03 | 2.34E-03 | 7.46E-04 | 3.92E-05 | 6.52E-04 | 1.40E-03 | 7.00E-04 | 1.25E-03 |
| VT | 1.76E-05 | 1.01E-05 | 6.14E-03 | 9.17E-03 | 5.71E-03 | 8.11E-03 | 2.62E-04 | 1.42E-05 | 1.81E-04 | 1.87E-04 | 1.76E-04 | 1.90E-04 |
| VA | 1.00E-02 | 3.48E-04 | 4.93E-03 | 5.34E-03 | 4.91E-03 | 5.77E-03 | 5.72E-03 | 3.10E-04 | 2.78E-03 | 3.17E-03 | 2.82E-03 | 3.07E-03 |
| WA | 1.34E-04 | 4.44E-05 | 3.99E-03 | 1.91E-03 | 4.33E-03 | 1.55E-03 | 5.39E-04 | 4.61E-05 | 1.03E-03 | 1.49E-03 | 1.01E-03 | 1.36E-03 |
| WV | 6.87E-03 | 2.53E-04 | 3.45E-03 | 1.58E-03 | 3.49E-03 | 1.55E-03 | 1.87E-03 | 2.17E-04 | 1.14E-03 | 1.67E-03 | 1.25E-03 | 1.59E-03 |
| WI | 1.95E-04 | 3.29E-04 | 4.29E-03 | 5.35E-03 | 4.28E-03 | 6.14E-03 | 8.87E-04 | 1.06E-04 | 1.33E-03 | 9.62E-04 | 1.59E-03 | 1.06E-03 |
| WY | 9.25E-02 | 1.07E-04 | 4.01E-03 | 2.61E-03 | 4.04E-03 | 2.22E-03 | 5.18E-03 | 5.35E-05 | 9.70E-04 | 1.45E-03 | 1.09E-03 | 1.37E-03 |

**Supplementary Table S7. COVID-19 hospitalization sd and hsd model fit of the peak of sunrise effect ($s_0$)**

| | η=0.026 | | η=0.05 | |
|---|---|---|---|---|
| | **sd** | **hsd** | **sd** | **hsd** |
| Pacific | 0.4550 | 0.4656 | 0.4602 | 0.4030 |
| East South Central | 0.4712 | 0.4694 | 0.4662 | 0.4094 |
| West South Central | 0.4852 | 0.4861 | 0.4647 | 0.4004 |
| Mountain | 0.4780 | 0.4827 | 0.4894 | 0.4387 |
| New England | 0.6246 | 0.5167 | 0.6680 | 0.4450 |
| South Atlantic | 0.4864 | 0.4880 | 0.4837 | 0.4219 |
| West North Central | 0.4750 | 0.4872 | 0.6814 | 0.4790 |
| East North Central | 0.5055 | 0.5113 | 0.5526 | 0.5162 |
| Middle Atlantic | 0.6053 | 0.5685 | 0.5795 | 0.5507 |

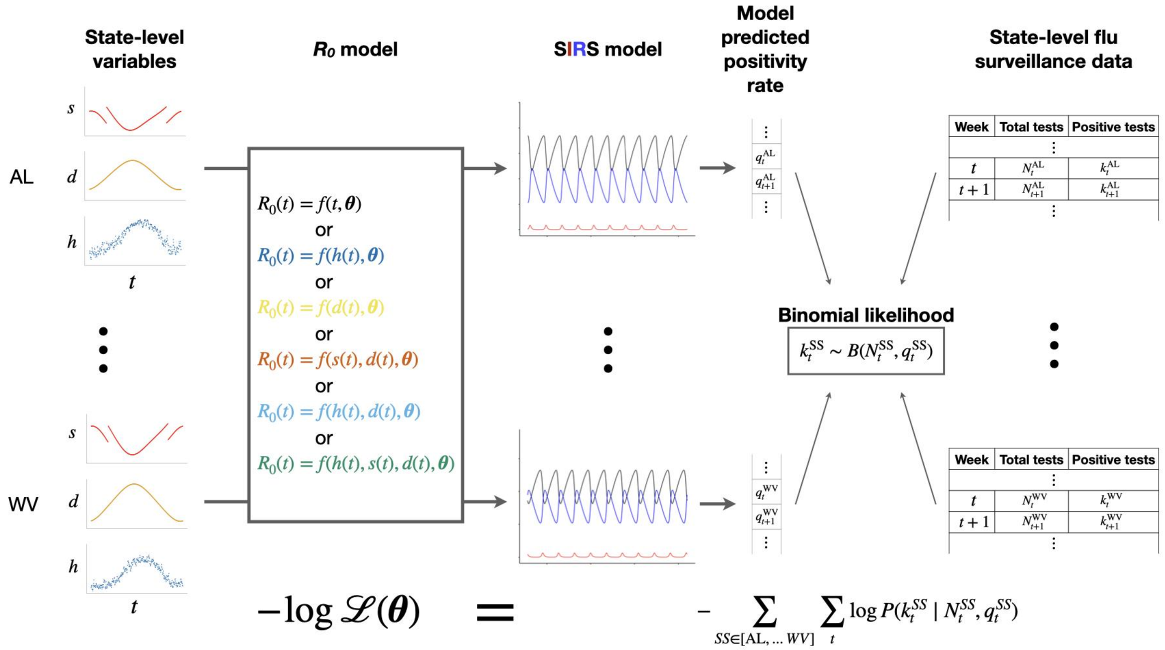


**Supplementary Figure S1. Evaluating goodness-of-fit of $R_0$ models using flu surveillance data of U.S. states.** Different $R_0$ models define the transformation from state-specific variables such as humidity ($h$), day length ($d$) and/or sunrise time ($s$) throughout the year to $R_0$ values, governed by a set of parameters $\boldsymbol{\theta}$ shared among all states. A separate instance of the SIRS model for each state predicts the infection rate and consequently the test positivity rate (see **Supplementary Materials**) given the $R_0$ values. Finally, the log likelihood of the model and parameter set $\boldsymbol{\theta}$ is calculated as the sum of log binomial probabilities of the observed weekly testing data over states. The log likelihood of each $R_0$ model is maximized for model comparison using Bayesian Information Criterion (BIC).

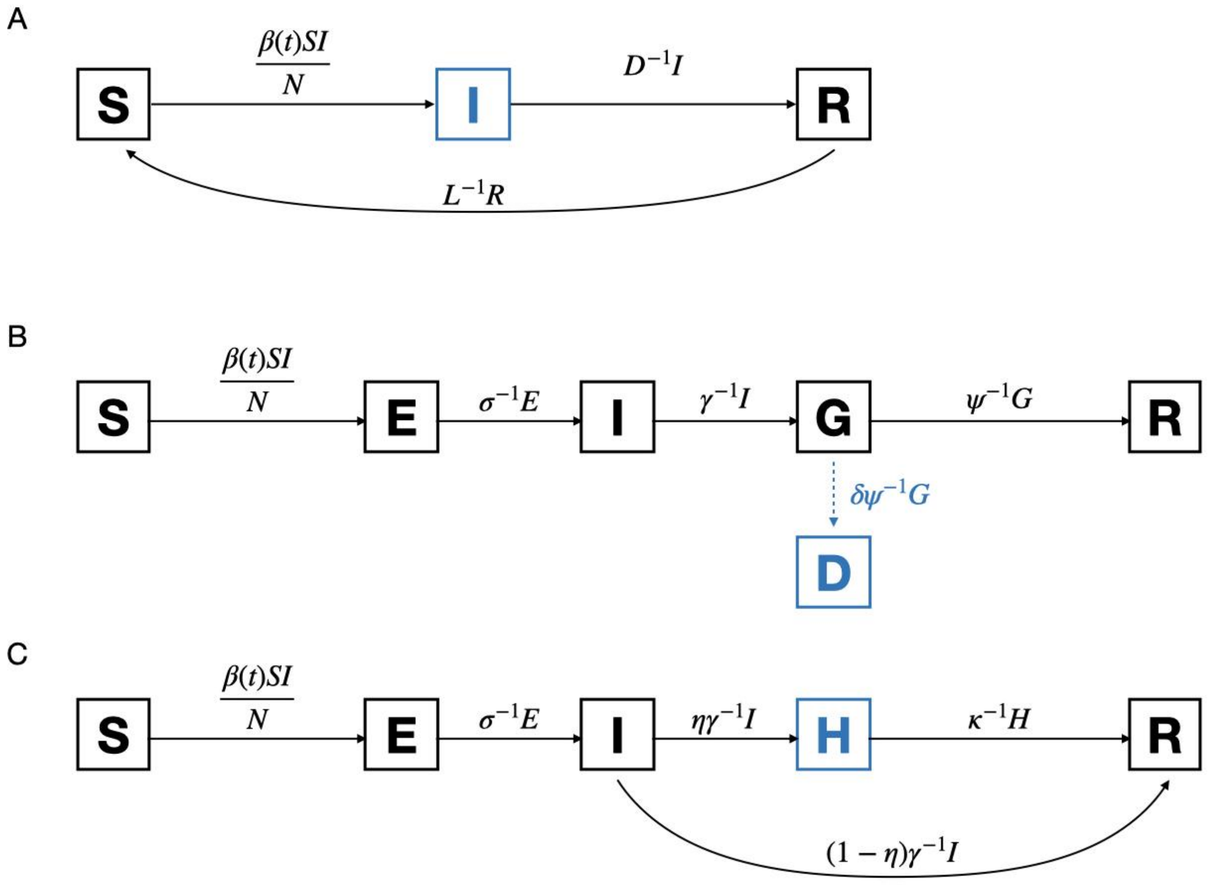


**Supplementary Figure S2. Three SIR-derived compartmental models** for modeling infection rate of endemic flu **(A)**, daily new deaths of COVID-19 **(B)**, and current hospitalization count of COVID-19 **(C).** See **Supplementary Materials** for definitions of the model parameters. Note that blue indicates the prediction of the model for which its likelihood is calculated. The dashed arrow in **(B)** emphasizes that the daily death count is treated as a sample of the resolving (G) compartment, and this sampling is independent of the dynamics of the model (i.e. cases resolved in both recoveries and fatalities enter the R compartment).

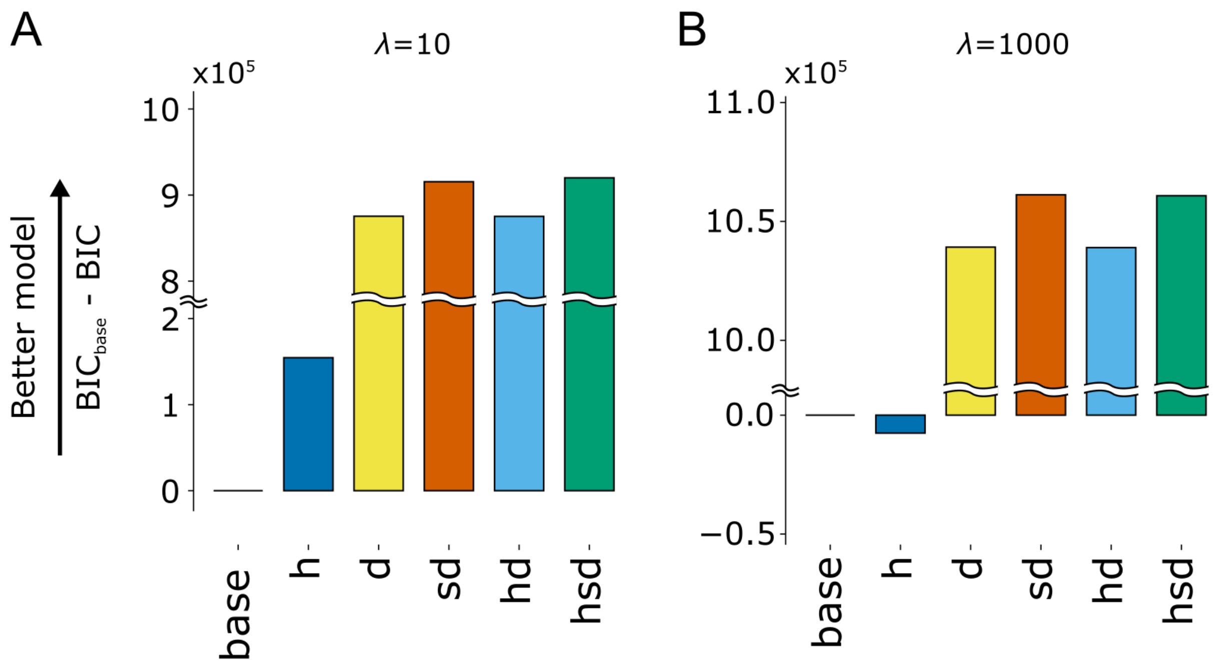


**Supplementary Figure S3. $R_0$ model fit for influenza surveillance data of 40 U.S. states from the 2010-11 season to the 2019-20 season.** Model fit is measured as the BIC for the base model minus the BIC for the base, climate (h), day-length (d), sunrise/day-length (sd), climate/day-length (hd) and climate/sunrise/day-length (hsd) models. Relative BIC values are shown for model likelihoods maximized under $\lambda$=10 (**A**) and $\lambda$=1000 (**B**).

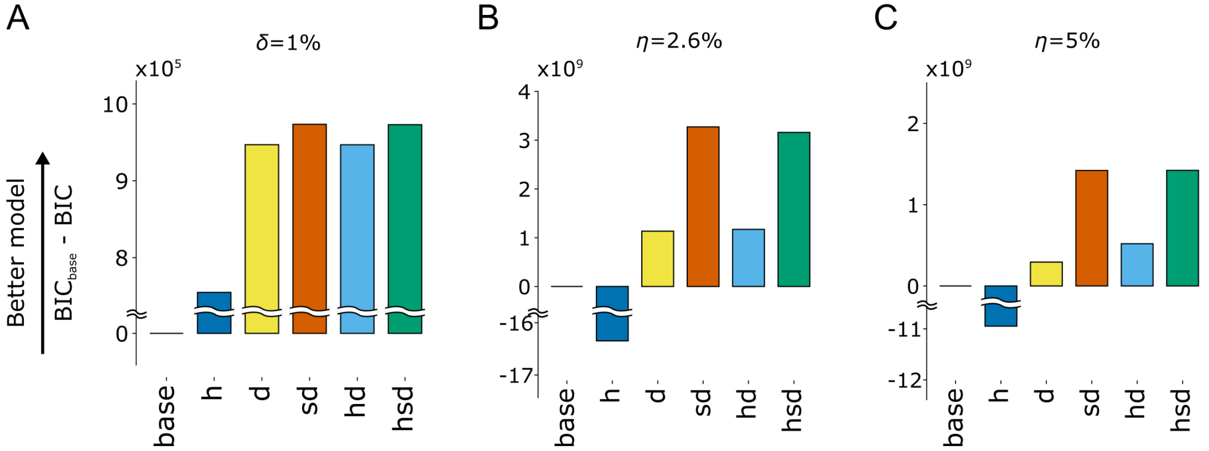


**Supplementary Figure S4**. ***$R_0$* model fit for state-level COVID-19 data in the United States.** Relative BIC values are shown for death models under a death rate of 1% **(A)**, as well as hospitalization models under hospitalization rates of 2.6% **(B)** and 5% **(C)**. Figure layout and description are otherwise similar to Figure S3.

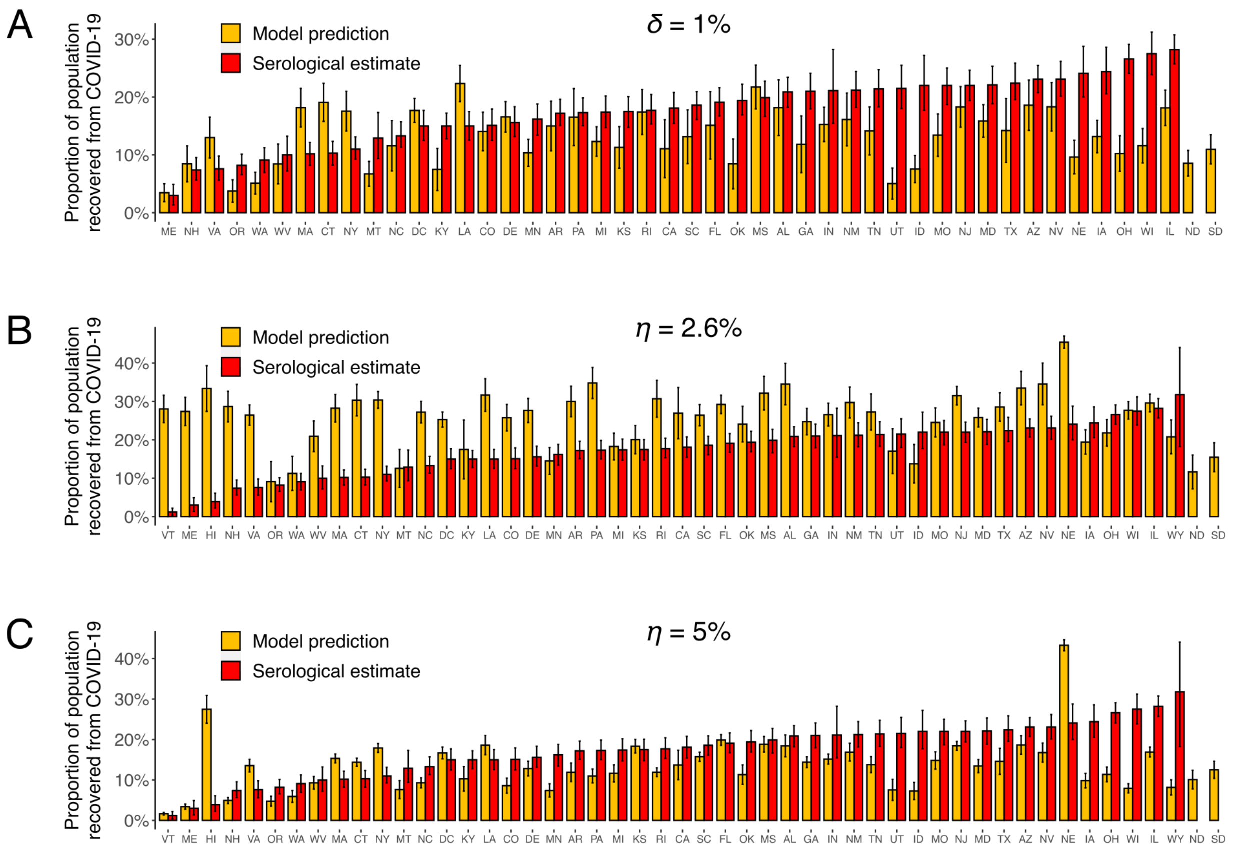


**Supplementary Figure S5. Comparison of the proportion of population recovered from COVID-19 by Jan 2021 inferred by the sunrise/day-length (sd) models and that estimated from serological surveillance**[14,15]. The proportion of the recovered population was inferred with the death model assuming a death rate of 1% **(A)**, as well as hospitalization models assuming a hospitalization rate of 2.6% **(B)** or 5% **(C)**. Error bars of model predictions indicate one standard deviation based on bootstrap replicates, whereas those of serological estimates represent confidence intervals as specified in ref [15].

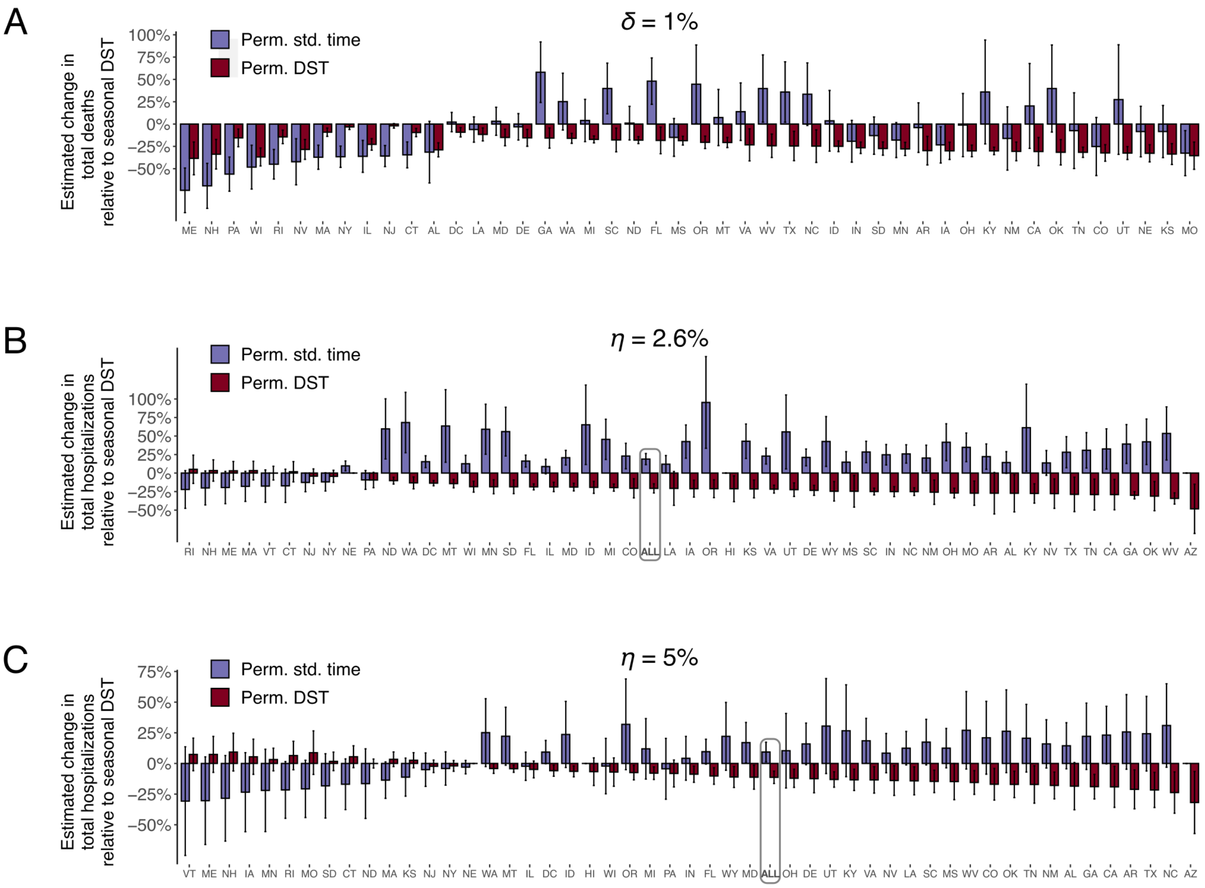


**Supplementary Figure S6. Estimated changes in the total number of COVID-19 deaths (A) and hospitalizations (B & C) in 2020 under alternative time policies.** Error bars indicate one standard deviation of simulation outcomes under bootstrapped model parameters.

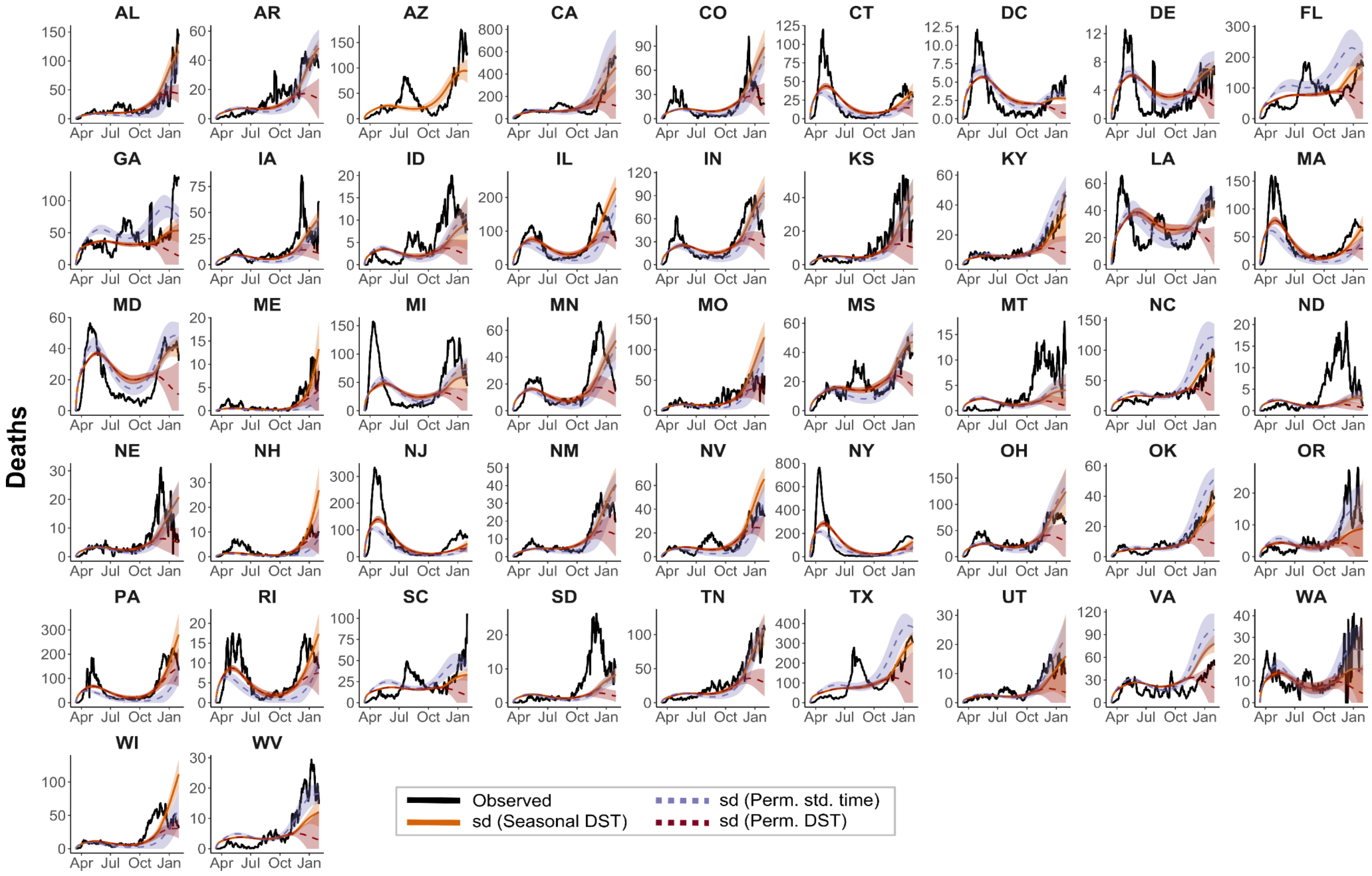


**Supplementary Figure S7. State level COVID-19 death data (from 03/13/2020 to 01/31/2021), the sunrise/day-length (sd) model fit and simulations of the sd model under alternative scenarios of DST observation.** The MLE of model parameters were obtained under a death rate (δ) of 1%. Figure descriptions are otherwise similar to Fig. 3C-F.

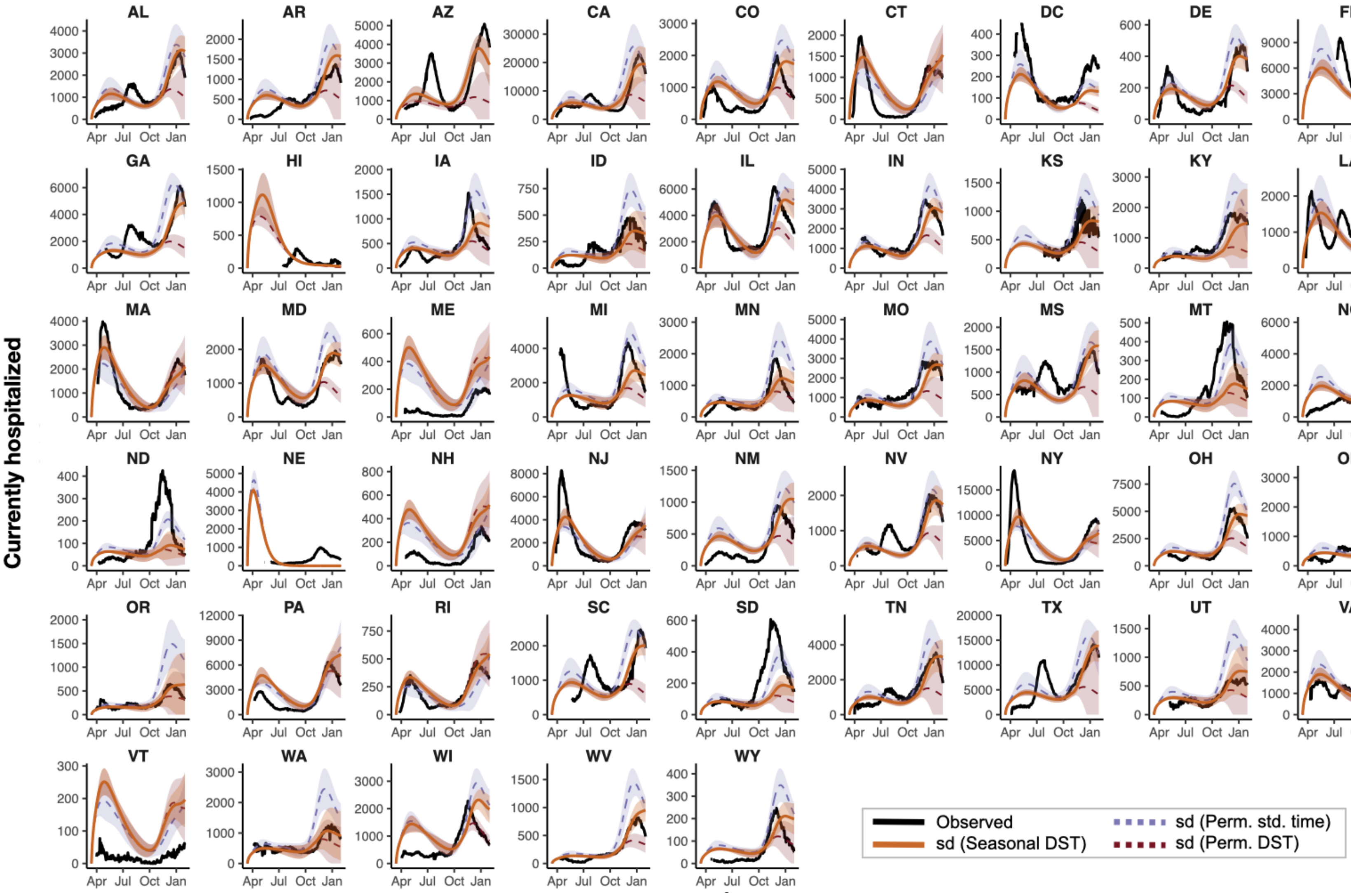


**Supplementary Figure S8. State level COVID-19 hospitalization data (from 03/13/2020 to 01/31/2021), the sunrise/day-length (sd) model fit and simulations of the sd model under alternative scenarios of DST observation.** The MLE of model parameters were obtained under a hospitalization rate (η) of 2.6%. Figure descriptions are otherwise similar to Fig. 3C-F.

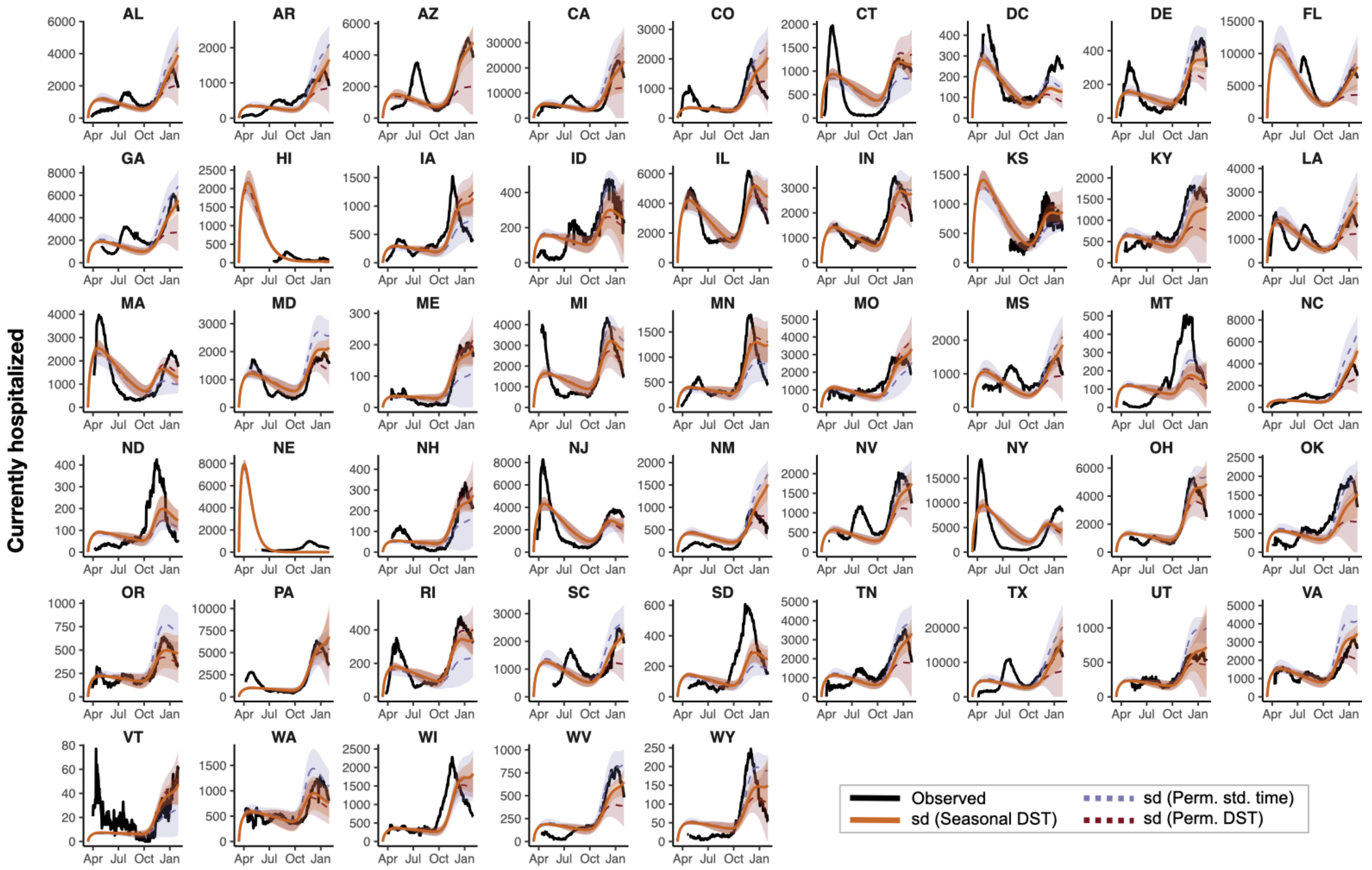


**Supplementary Figure S9. State level COVID-19 hospitalization data (from 03/13/2020 to 01/31/2021), the sunrise/day-length (sd) model fit and simulations of the sd model under alternative scenarios of DST observation.** The MLE of model parameters were obtained under a death rate (η) of 5%. Figure descriptions are otherwise similar to Fig. 3C-F.

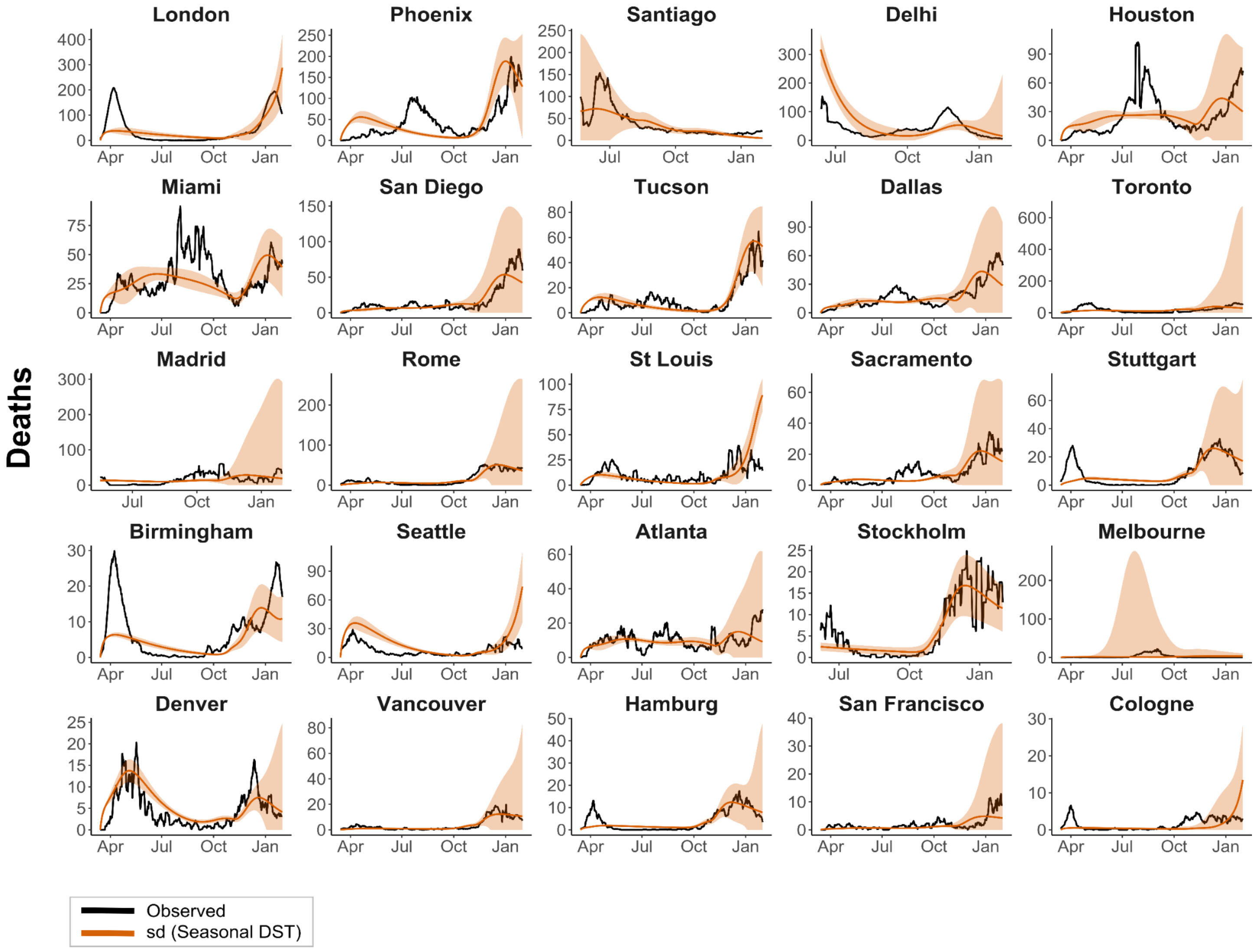


**Supplementary Figure S10. COVID-19 death data in 25 urban centers and the sunrise/day length (sd) model fit.** Figure descriptions are otherwise similar to Fig. 4.

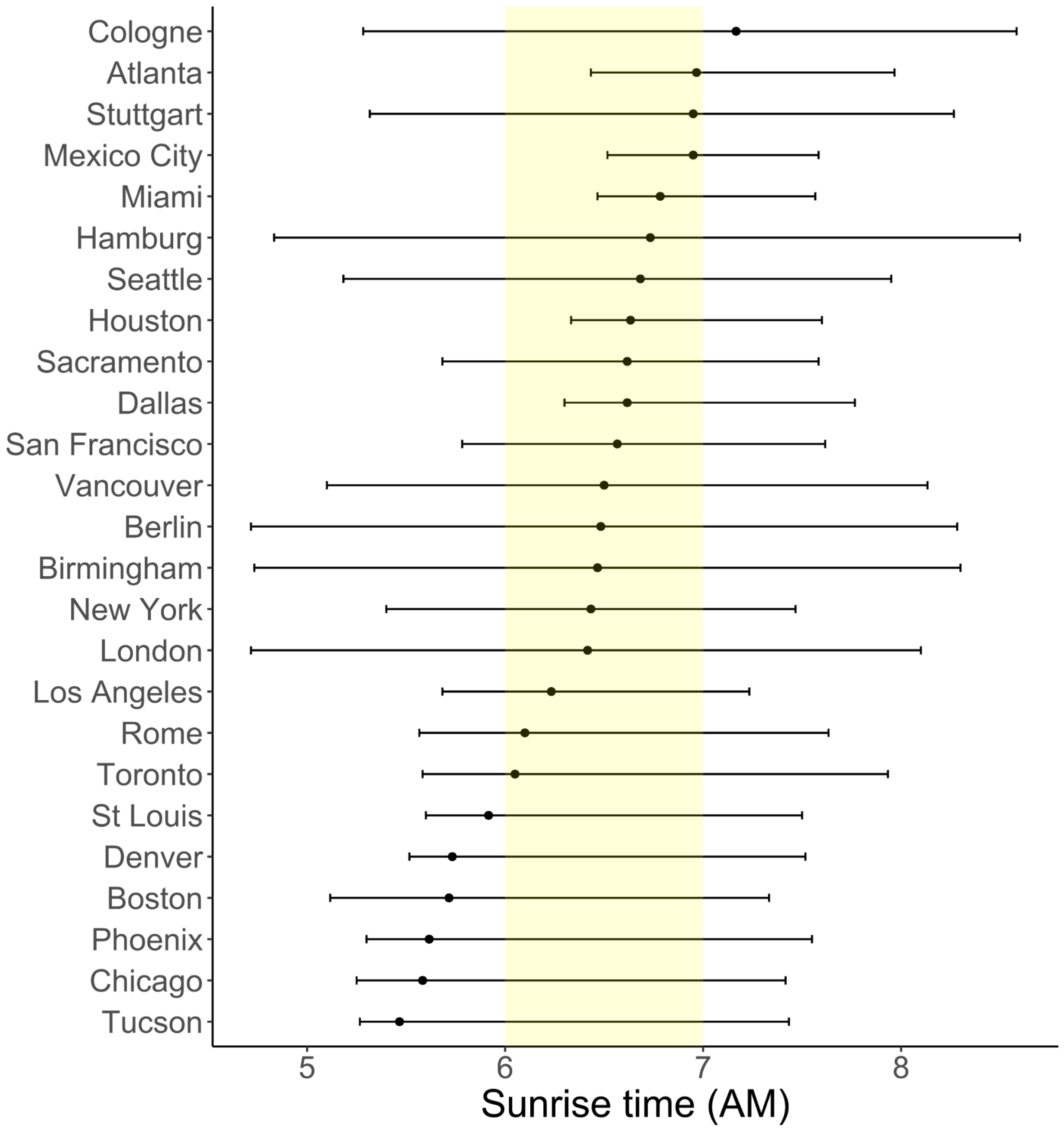


**Supplementary Figure S11. Sunrise time in 25 cities at the estimated peak of daily infections in the first wave of COVID-19.** Time points are based on peak observed deaths in each city between January 1., 2020 and October 1., 2020, with the time lag between peak deaths and peak infections assumed to be 10 days. Bars indicate the range of possible sunrise times in each city across the year. The vertical yellow bar shows the period of likely susceptibility between sunrise times of 6am and 7am. Six cities from our full set were excluded: Santiago, Delhi, Stockholm and Madrid were excluded because of missing data in early 2020 and Melbourne and San Diego were excluded because these cities did not experience a first wave during the time period analysed.

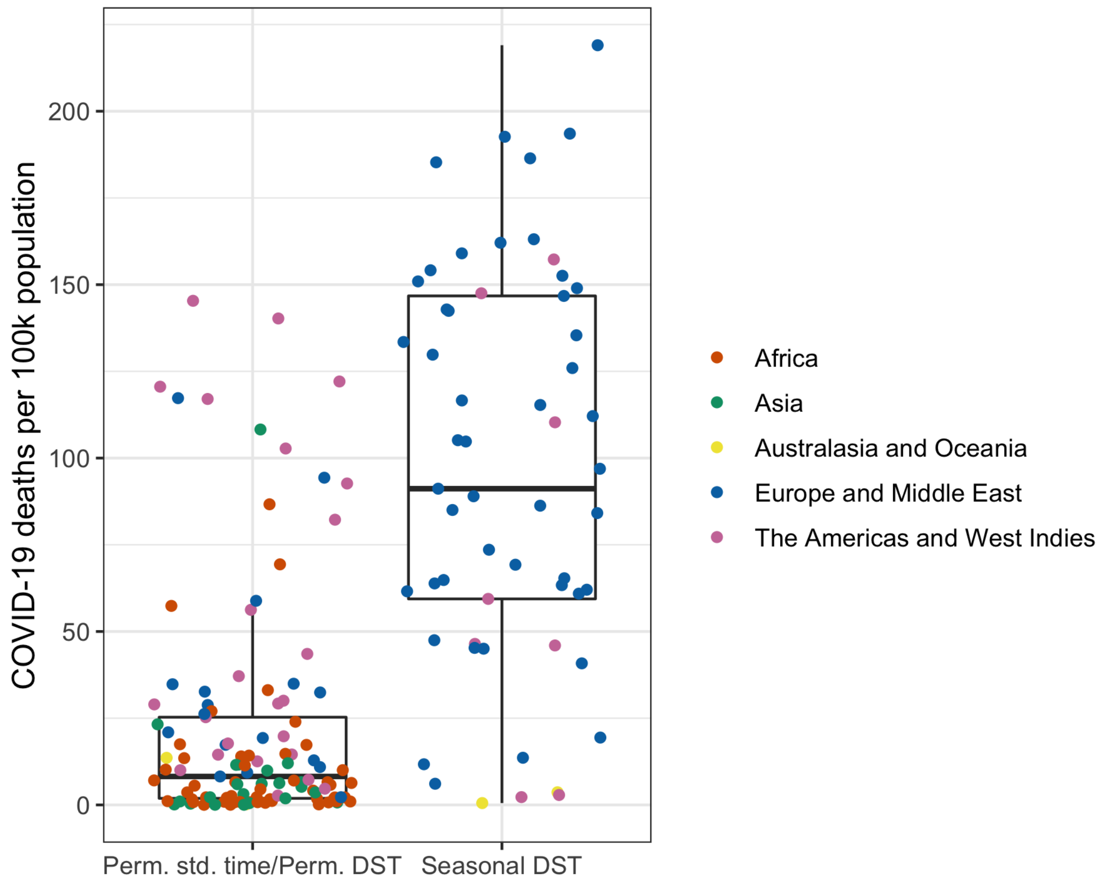


**Supplementary Figure S12. COVID-19 mortality rate per 100,000 population in countries with or without seasonal DST.** Asian and African countries without seasonal DST have lower mortality rates than in countries that do observe clock changes.
